# Stakeholder perspectives on contributors to delayed and inaccurate diagnosis of cardiovascular disease: a UK-based qualitative study

**DOI:** 10.1101/2023.09.28.23295847

**Authors:** K Abdullayev, O Gorvett, A Sochiera, L Laidlaw, TJA Chico, M Manktelow, O Buckley, J Condell, RJ Van Arkel, V Diaz-Zuccarini, Faith Matcham

## Abstract

**Objective:** The aim of this study is to understand stakeholder experiences of cardiovascular disease (CVD) diagnosis to support the development of technological solutions that meet current needs. Specifically, we aimed to identify challenges faced by stakeholders in the process of diagnosis of CVD; to identify discrepancies between patient and clinician experiences of CVD diagnosis, and to make recommendations for the requirements of future health technology solutions intended to improve CVD diagnosis.

**Design:** The qualitative data was obtained using semi-structured focus groups and 1-1 interviews.

**Participants:** UK-based individuals (N = 32) with lived experience of diagnosis of CVD (n = 23) and clinicians with experience in diagnosing CVD (n = 9).

**Results:** Thematic analysis of focus groups and interview transcripts produced four key themes related to challenges contributing to delayed or inaccurate diagnosis of CVD: Symptom Interpretation, Patient Characteristics, Patient-Clinician Interactions, and Systemic Challenges. Sub-themes from each theme are discussed in depth.

**Conclusions:** Challenges related to time and communication were greatest for both stakeholder groups, however there were differences in other areas, for example patient experiences highlighted difficulties with the psychological aspects of diagnosis and interpreting ambiguous symptoms, while clinicians emphasised the role of individual patient differences and the lack of rapport in contributing to delays or inaccurate diagnosis. Key takeaways from this qualitative study were summarised into a table of considerations to highlight key areas that require prioritisation for future research aiming to improve the efficiency and accuracy of CVD diagnosis using digital technologies.

## INTRODUCTION

Approximately 6.8 million people have heart disease in England, with 25% of deaths in the United Kingdom (UK) caused by cardiovascular disease (CVD) (1). The economic and social burden of CVD continue to increase globally (2). CVDs cost the UK economy an estimated £7.4 billion annually, rising to £15.8 billion when considering wider economic costs (3). CVDs contribute to high excess primary and secondary care costs, which are doubled when patients have previously suffered cardiovascular events (4). There are also severe psychological and social consequences of diagnosis which extend beyond the patient to their families and support network (5,6).

Despite the impact of CVD on the UK economy and population, in the years prior to CVD diagnosis, approximately 50% of patients had a primary care consultation and only 24% of patients following the recommended pathway to diagnosis (7). Moreover, a recent systematic review revealed misdiagnosis of heart failure ranges from 16% in patients discharged from hospital to 68% in primary care services (8). Delayed and inaccurate diagnoses have also been found to be common in specific CVDs, such as transthyretin amyloid cardiomyopathy (9) and pulmonary embolism (10). Delayed or misdiagnosis contribute to patients receiving inappropriate treatments or undergoing unnecessary evaluations (11,12). Adverse outcomes may result from effective treatment not being received until the disease is more advanced (13). A missed diagnosis of congestive heart failure is associated with increased hospital readmission rates (14), and with a two-fold increased risk of death (15). Avoiding misdiagnosis and reducing diagnosis delays are critical for improving patient outcomes and reducing healthcare costs.

As a result of increased pressures on healthcare systems worldwide, accelerated by the recent COVID-19 pandemic, there is growing interest in the potential for technology-based solutions to improve healthcare delivery. Recurring lockdowns prompted a sharp rise in the use of remote measurement technologies in healthcare (16–18). Digital Twins (DTs) are an example of an emerging technological solution for challenges faced in the diagnosis of CVD (19–21). DTs use live data to build a digital representation of patient molecular, physiological and lifestyle status over time (22). DTs can be used to predict future trends (21) and personalise treatment plans (19). Although the potential of DT technology has yet to translate into clinical care pathways (22–24), the shift towards increased remote care during and following the COVID-19 pandemic, has paved the way for greater technological innovation.

A recent review of telehealth use during the COVID-19 pandemic outlined lack of human contact in care, confidentiality, data security, and accessibility and training in the use of new platforms as key challenges associated with the implementation of technology into healthcare (18). Healthcare technologies need to consider how these challenges impact patient and clinician engagement and more work is needed improve our understanding of user experience to produce sustainable improvements in CVD diagnosis which can be readily implemented into clinical care. Existing work has shown the critical requirements of a digital technology platform for the self-management of CVD (25); however, the point of self-management can only be reached once an accurate and timely diagnosis has been made. Technology plays an important role in facilitating this process.

The aim of the present study is to develop a deeper understanding of stakeholder experiences of CVD diagnosis, to support development of technological solutions which meet current needs. Specifically, our objectives to:

1. Identify current challenges faced by stakeholders in diagnosis for CVD.
2. Identify discrepancies between patient and clinician experiences of CVD diagnosis.
3. Make recommendations for requirements of future health technology solutions for improving CVD diagnosis.

## METHODS AND MATERIALS

Our methodology and procedure is outlined in detail in a published protocol (26). The study was conducted and reported according to Consolidated criteria for Reporting Qualitative research (COREQ; (27)) guidelines.

### Study Design

We used semi-structured focus groups in people living with CVD to generate discussions of shared experiences during their diagnosis journey. We additionally conducted 1:1 interviews with clinicians to increase our flexibility around their schedules and collect information across a range of different clinical specialities.

This study was reviewed and approved by the Sciences & Technology Cross-School Research Ethics Council at the University of Sussex (reference ER/FM409/1).

### Patient and Public Involvement

Prior to starting recruitment, all participant-facing materials were reviewed by a Sheffield-based cardiovascular patient advisory group. This ensured all language used in the information sheet, consent form, and focus group topic guide was accessible and easy to understand.

### Study Population

Figure 1 shows a flowchart of participant recruitment. Inclusion criteria for lived experience individuals were: a previous diagnosis of CVD; aged 18 or over; able to speak English sufficiently for participation; able to consent to participation. Exclusion criteria included: major cognitive impairment or dementia preventing participation. The inclusion criteria for clinicians were: > 6 months of experience in diagnosing patients with CVD; aged ≥18 years; able to speak English; and able to consent to participation.

**Figure 1.**
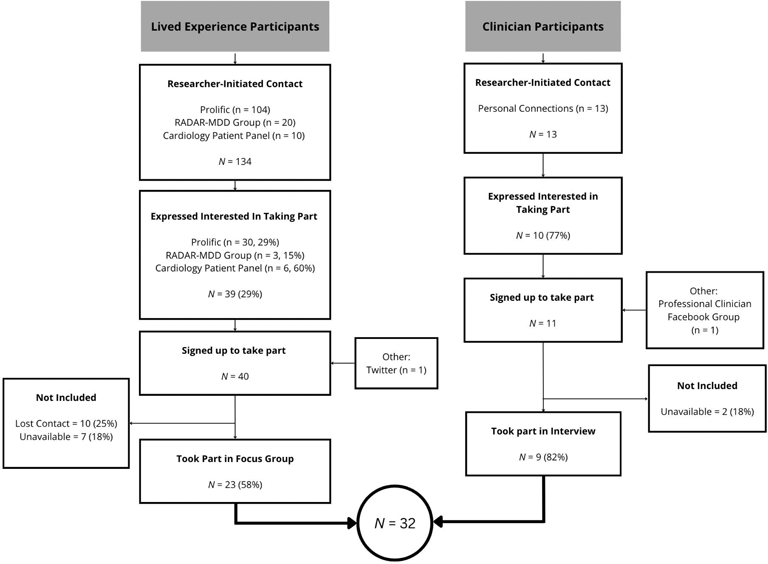

### Procedure

Lived experience participants were recruited using convenience sampling via: Prolific; a cardiology patient group; participants from a previous research study who had consented to be contacted for future research purposes (28). Study details were also shared on Twitter. Individuals interested in participating were contacted via email to arrange an introductory phone call to confirm interest and eligibility. In this meeting, FM described the research and the procedure of the study.

Clinicians were recruited using purposive sampling via personal and professional connections, and a registered GP Facebook group. All information was given to clinicians via email prior to the online interview.

Consent and baseline demographic data were collected via online Qualtrics surveys prior to qualitative data collection (Appendix 1). The focus groups and interviews follow a pre-approved, semi-structured question schedule, split into two sections (Appendix 2). All focus groups and interviews were conducted online over Zoom, with focus groups lasting about 90 mins and interviews ranging between 30-90 minutes, based on clinician time availability. Interviews and focus groups were facilitated by KA; a female Psychology graduate, working full-time on the project. KA had no ongoing relationship with the participants and was not involved in their clinical care. She had neither prior experience in cardiology nor assumptions or expectations of the data. Data collection was supported by two research facilitators (OG and AS) who made field notes during interviews and focus groups. Field notes were destroyed once transcripts were deidentified and finalised.

The focus group and interviews were audio-recorded, anonymised then transcribed verbatim prior to analysis. Transcripts were validated by KA, OG, AS to confirm transcript accuracy.

### DATA ANALYSIS

Data relating to patient and clinician experiences of the CVD diagnosis pathway were included in the current analysis. Sample sociodemographic characteristics were described, alongside Depression, Anxiety and Stress scores (DASS, (29)) to understand underlying levels of depression, stress and anxiety at the time of participation. Overall scores were classified into three severity groups – Normal (Subclinical), Moderate, and Severe – based on validated thresholds (30).

The present analysis adopted a phenomenological approach using reflexive inductive thematic analysis. KA used NVivo to conduct the analysis, following the steps recommended by Braun and Clarke (31). This involved: data familiarisation; initial code generation; theme identification; collaborative theme review; and final definition and naming of themes. Coding was initially conducted by KA, with secondary coding and review conducted by OG and AS to validate theme extraction. The findings were discussed between each of the reviewers before a final decision was made regarding themes and sub-themes that would be reported.

For the last stage of analysis, we collaborated with an experienced patient and public contributor, who has experience of conducting qualitative research (LL) to create an additional layer of validation for our framework. This involved meeting over Zoom to discuss our coding framework and reinterpreting the data to accurately reflect a patient perspective. In line with recent developments in the literature (32), saturation was not assessed as ideas of thematic saturation are not consistent with the values and assumptions of reflexive thematic analysis.

### Scientific Rigour

To increase scientific rigour of our findings, the results of the first round of thematic analysis were presented to clinicians in the form of a research poster at the British Cardiology Society conference to increase transferability of our results to a wider sample. We also consulted with the London-based NIHR Maudsley Biomedical Research Centre’s Race, Ethnicity and Diversity (READ) advisory group to provide further cultural insight on our preliminary findings, which were presented via a series of presentation slides summarising the key findings so far.

## RESULTS

### Sample Demographics

A total of four focus groups (N=23) and nine interviews were conducted with a total of 32 individuals contributing data to the study. The total number of participants represents 63% of interested individuals and 22% of individuals initially contacted. Table 1 summarises the demographic and clinical characteristics of the sample.

**Table 1.**
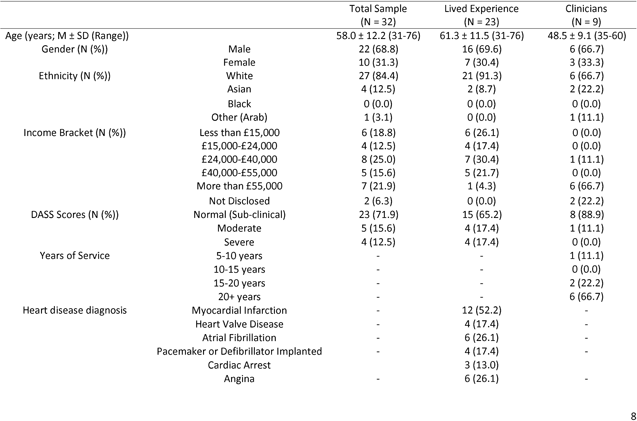
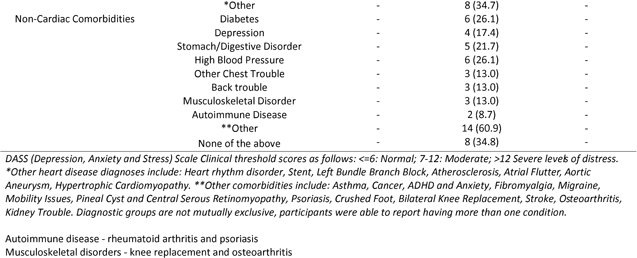
Demographics of the Sample (N = 32)

Four overarching themes and 34 subthemes were identified (Figure 2) and quotes for each subtheme are presented in Appendix 3. The 4 major themes were: Symptom Interpretation, Patient Characteristics, Patient-Clinician Interactions, and Systemic Challenges.

**Figure 2.**
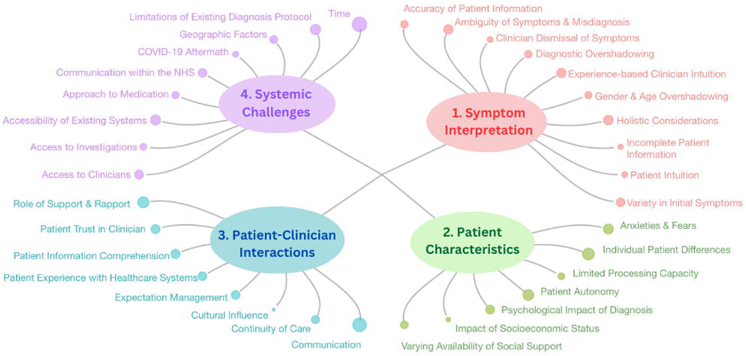

#### Theme 1: Symptom Interpretation

Data revealed the wide variety of symptoms experienced prior to diagnosis – from “sweating” and “tachycardia” to “swollen legs” and “severe acid reflux” – highlighting the diversity in experiences between patients with different conditions, ranging from atrial fibrillation to myocardial infarction. From the patient perspective, the ambiguity of symptoms they were experiencing added to confusion about whether to reach out to their doctor: “is this AF or am I just getting myself worked up with it? So, you kind of doubt your reliability, don’t you?” (P18).

Difficulty interpreting the cause or severity of their symptoms negatively affected their ability to express themselves in appointments with a clinician: “it really concerns me then, when I’m sat in front of the person who needs to know what has been happening that I can’t articulate it very accurately.” (P17). Similarly, clinicians describe how patient interpretation of their own condition can affect their ability to efficiently diagnose them and how their personal biases affect the information patients choose to share, for example when it comes to alcohol, there is a “natural will to reduce down the amount that they’re saying that they’re actually drinking” (CL10). This can result in incomplete information being passed on to the clinician, preventing informed decisions regarding their diagnosis. Although clinician perspectives seemed to prioritise “looking at patient background, and their socio-economic background, their personal family history…smoking as well, diet, stress, exercise, basically the whole picture” (CL12), patients had mixed experiences when it came to holistic considerations of their health condition: “their job seems to be more physical in terms of, you know, treating the condition rather than, you know, the mental aspect of it or the ongoing aspects of it” (P29).

Patients also described challenges related to diagnostic overshadowing: “whenever I I went to my GP and said, I’m not feeling well, he would say, well, you have a heart problem but I’m pretty sure that the last two years I’ve been suffering from something other than my heart, or in addition to my heart.” (P9). This problem was echoed by clinicians, who acknowledged that “chest cardiac symptoms can sometimes be very vague and overlap with other diagnoses” (CL9), contributing to false attribution of non-cardiac symptoms to a cardiovascular disease. Demographic factors, such as age and gender, also created difficulties with symptom interpretation: “One of the consultants told me that I don’t look like a heart problem because I am still young, and I don’t, I’m not overweight, or I don’t drink or alcohol anything like that. So, I shouldn’t have a heart problem basically” (P13), highlighting how members of groups less likely to suffer from heart disease might have less emphasis placed on their relevant symptoms and experiences.

Both clinicians and patients described experiences related to intuition and previous experience guiding decisions regarding their symptom interpretation, with one patient sharing how they “just woke up and knew something wasn’t right and when the ambulance came and they said, what’s wrong? And I said, I don’t know but something is” (P20) and a clinician describing how they use their “knowledge of the biology, physiology, pharmacology, pathology, and histopathology, and as well as clinical knowledge” (CL1) when taking a patient history.

This theme provides insight into the importance of supporting both patients and clinicians in interpreting systemic and overlapping symptoms linked to CVD.

#### Theme 2: Patient Characteristics

Focus group discussions revealed the vast array of individual differences amongst patients which influence their personal experience of being diagnosed with CVD, with one patient nicely summarising how “everyone is totally different, as is what they want, what they want to know, how much they want to know, and how they want to know it” (P4). These differences manifested in several ways, for example in the level of autonomy with which they managed their condition, with one patient describing how their GP stopped “following up, asking for my readings, so I just stopped doing it.” (P10), while another patient “just took myself to the GP” (P28). Meanwhile, clinicians seem to expect quite a high level of autonomy from patients, as ‘there’s no active monitoring. The monitoring itself would depend on the patient staying in touch with me.” (CL12). There were differences in how much support patients had during their diagnosis with some reflecting how “they owe a lot of people a lot” (P10) for their support, while others said their “greatest challenge was I live on my own… and there was no support” (P9). Clinicians also discussed how differences in socioeconomic status will impact diagnosis and patient care, as lower literacy levels require more support, with one clinician describing how they “bring them in, and I’ve got forms and sheets that I go through with them and give them information” (CL10).

There was also substantial evidence for the psychological impact of CVD diagnosis, with one patient describing how their “life seemed to stand still. Not just physically, but also mentally” (P5,) and how there was “a lot of anxiety around the condition, particularly whilst or once I was diagnosed…that was getting really quite troublesome” (P17). Similarly, clinicians brought up similar issues regarding fear and anxiety management, reflecting on the importance of “trying to remove the fear from the from the patient trying to de-escalate them” (CL12) and considering patient mental processing capacity during the time of diagnosis and how this might lead patients to “delete… and distort a lot of the information” (CL10).

This theme provides insight into how healthcare services and procedures could improve quality of care and patient outcomes by considering patients as individuals, with different needs and expectations.

#### Theme 3: Patient-Clinician Factors

Focus group discussions revealed frustrations from some patients regarding the lack of communication and expectation management – leaving them “initially a bit confused” (P25) and unsure of “what the future was going to hold or how long I was going to survive” (P29) – and the lack of continuity of care, as “now if you… phone to speak to a doctor, you invariably get somebody you’ve never spoken to before” (P22). Clinicians expressed how time affects communication. They are “only given 10/15 minutes to see one patient, which I don’t think is enough” (CL9) and awareness of lack of continuity of care means that “particularly in our older clientele…they feel abandoned” (CL10).

Meanwhile, other patients relayed the opposite sentiment, providing positive stories of how “the cardiologist… certainly did a lot to save my family from me dying” (P2) and a general sentiment of trust in clinician competence: “you get to realize that they know 100 percent more than you do so it’s common sense to accept what they are saying.” (P26). Clinicians also described techniques they use to help patients understand their diagnosis, “like drawings, pictures, things that would be able… to inform the patient better about the condition” (CL7).

There was an element of luck in the type of experience patients had, with some patients describing how lucky they were to have been “seen by a very astute newly qualified GP” (P28) while others felt their doctors “were guessing much of the time” (P9). Clinicians seemed to be aware of this, stating that “depending on the consultant they get, they will feel very informed and very supported, or they will equally feel very judged and dismissed” (CL10). Many of the challenges related to patient-clinician interactions were described by clinicians as being out of the hands of GPs and cardiologists, as they were the result of systemic protocols and not individual decisions, for example one clinician recalls how she doesn’t “start drugs going into a bank holiday, because I know the out of hours doesn’t exist” (CL10). Nonetheless, interview data suggested clinicians played an active role in mitigating the impact of existing systemic limitations and creating a relationship based on trust and open communication, as they emphasise that “the patient needs…to be willing to work with the clinicians and if the patient refuses to work with the clinician then no matter how hard the clinicians try you know it’s not going to work” (CL7).

This theme provides insight into the importance of building meaningful relationships between patients and clinicians for improving patient outcomes and indicates areas where greater standardisation of care may be of benefit.

#### Theme 4: Systemic Challenges

Many systemic issues affected both patients and clinicians, such as “the complete lack of communication between the medical staff, the nurses in this case and the senior doctors and…surgeon” (P19) and the fact that “ED has no mechanism to give [GPs] feedback” (CL10). Both groups also faced obstacles resulting from existing protocol, such patients having to go “through a kind of interrogation to find out how serious” (P8) their condition was to be able to book an appointment, or clinicians who “feel very strongly that this is a ischaemic heart disease…but if it doesn’t meet the criteria, and then it is rejected” (CL12). In addition to the limitations of existing protocols, there are several resource-related challenges that are preventing patients from receiving an accurate and efficient diagnosis, including access to clinicians, access to investigations and time-related difficulties. Both patients and clinicians describe difficulties getting appointments, with one patient having “only seen the cardiologist once in two years” (P9), while one clinician boldly stating that “we need to empower a patient as much as possible… because the NHS… can no longer provide that sort of mollycoddling” (CL8). Although, other clinicians did not share this sentiment and felt that “the poor patient is stick in the middle” (CL2) of issues related to the NHS and even though “what is lifesaving usually gets done…for life changing procedures…there is no capacity to see all these people so quickly” (CL1).

Geographic factors appeared to exacerbate these issues, as in isolated regions “the nearest cardiologist is…over several hundred miles away” (CL9) and there is “a real practical issue about getting access to tests” (CL8). There is some evidence to suggest that patients and clinicians feel previously existing systemic issues, particularly in relation to access to clinicians, has worsened as a result of the COVID-19 pandemic, as patients feel “you can’t get to see [the GP] obviously as easily” (P18) and clinicians are also aware that “in the kind of post-Covid era, patients have a lot of problems getting access to a GP so I mean, that’s a big barrier right now, and we have we have a limited resource” (CL13).

Finally, there were also patient concerns with the approach to medication within the healthcare system and a general lack of understanding in the way their medications were managed (or weren’t managed) by their clinicians, as “no one questions” repeat prescriptions (P5) or keeps track of the dosage until the patient questions it. Overall, our data showed how existing issues within the healthcare system interact with each other to ultimately disadvantage both patients and clinicians, and as a result, contribute to poorer outcomes for CVD patients.

This theme highlights how limited access to resources, in the form of clinicians, investigations, and time, act as barriers to efficient and accurate diagnosis of CVD.

Table 2 breaks down the size of each theme and subtheme, split by population group. Despite variation in subtheme sizes between the groups, the overall totals for each major theme are similar, suggesting that patients and clinicians face comparable challenges, despite some variation in more specific difficulties captured by the subthemes. The four major themes are relatively similar in size, although the Systemic Challenges theme was the largest and the Patient Characteristics theme was the smallest.

**Table 2.**
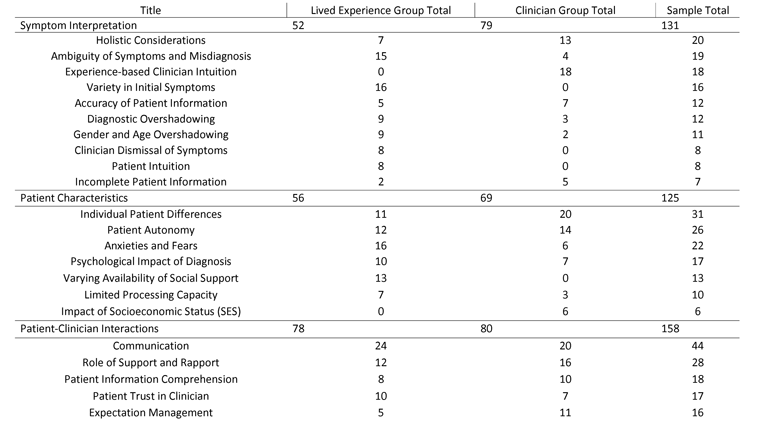
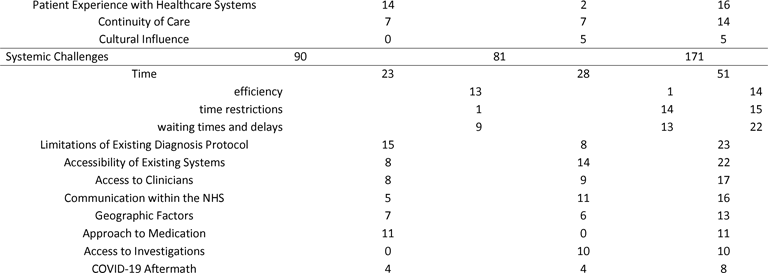
Breakdown of Theme and Subtheme Size.

### Scientific Rigour

Cross-validation of findings with clinicians attending a cardiology conference and members of a Race and Ethnicity advisory group resulted in no major changes in the interpretation of the data, increasing confidence in the translatability of the present findings.

## DISCUSSION

The results of this study suggest clinicians and patients face a variety of challenges preventing accurate and timely diagnosis of CVD. These difficulties were categorised by experiences related to Symptom Interpretation, Patient Characteristics, Patient-Clinician Interactions and Systemic Challenges. All four themes were relatively similar in size, though the largest was Systemic Challenges

Challenges related to ‘Time’, including time restrictions, long waiting times and delays, and efficiency, had the greatest number of references, consistent with existing knowledge of issues with time and resources within the NHS (33). The second biggest sub-theme was ‘Communication’ between patients and clinicians, including a combination of positive and negative patient experiences. This is consistent with a previous qualitative study including both patients and healthcare professionals, which highlighted how problems with communication lead to a lack of patient understanding of their CVD (34). A systematic review found that patient-clinician interactions influence patient capacity to engage in self-care for their heart condition via their ability to influence patient understanding of their condition (35). Thus, our study complements wider knowledge of the direct and indirect influence of quality communication on patient healthcare outcomes (36,37). The third largest sub-theme was related to ‘Individual Patient Differences’, highlighting the role of patient differences in determining both patient and clinician experiences during diagnosis. The growing interest in the implementation of personalised healthcare via wearable devices and digital medicine provides opportunity to account for these differences and improve patient experience and health outcomes (38,39).

There were also several smaller subthemes that were found in our data that are also supported by existing literature. For example, we found that location contributed to systemic challenges such as access to clinicians and investigations, consistent with previous studies investigating the variability in access to diagnostic tests (40) and inequity in GP supply (41) across the UK. Also, previous findings related to post-Covid-19 challenges in CVD diagnosis were supported by our data from both patient and clinician perspectives, although the smaller size of the subtheme may be due to many of our patient experiences being pre-Covid-19 and thus were not affected by delays following nationwide lockdowns (16,42). However, issues related to diagnostic overshadowing and misdiagnosis due to comorbidities were less substantial than expected, given existing knowledge of difficulties diagnosing vague symptoms and falsely attributing overlapping symptoms to pre-existing non-cardiac conditions (43,44). This may be due to clinicians not feeling comfortable admitting that they struggle to accurately diagnose their patients; meanwhile, ‘Ambiguity of Symptoms and Misdiagnosis’ came out as one of the largest subthemes from patient data, suggesting that difficulties with accurate symptom interpretation can prevent some patients from reaching out to healthcare professionals.

Our results shed light on the differences between patient and clinician experiences, highlighting the importance of considering how barriers to diagnosis may be affecting each group differently. The weight of evidence for the four major themes did not differ substantially between lived experience and clinician groups, however there were differences between the size of subthemes. Although ‘Time’ and ‘Communication’ were the largest subthemes for both participant groups, ‘Individual Patient Differences’, ‘Role of Support and Rapport’ and ‘Experience-based Clinician Intuition’ were the largest subthemes from clinician data, while ‘Anxieties and Fears’, ‘Ambiguity of Symptoms and Misdiagnosis’ and

‘Limitations of Existing Diagnosis Protocol’ were the largest subthemes from patient data. Notably, both participant groups acknowledged in some way the psychological aspects of CVD diagnosis, which is supported by existing findings showing moderate levels of depression and anxiety and significant life changes in people with heart failure (45). Nevertheless, patient data revealed more about the patient mental load associated with diagnosis, while clinicians shared their opinions and experiences related to providing support for patients, which is in line with previous research looking at the role of clinicians in providing relief and support following a cardiac event (46).

A summary of considerations arising from this research are listed in Table 3. These could be used for future research and development of health technology solutions aiming to improve accuracy and efficiency of CVD diagnoses and improve patient outcomes by reducing mortality and increasing treatment efficacy.

**Table 3.** Considerations for Future Research.

| Considerations |
| --- |
| - Improve efficiency of processes and interactions during diagnosis, to aid both clinicians and patients when time is limited<br>- Implement strategies to improve communication between patients and clinicians so patients feel informed throughout the process of their diagnosis and have access to accurate and appropriate information<br>- Acknowledge individual patient differences when implementing any solutions or new systems by allowing personalisation and flexibility in the use of the solution<br>- Incorporate support and rapport building between patients and clinicians in systems aimed to improve accuracy and efficiency<br>- Be conscious of the extent to which patients are expected to take autonomy over their care and how (in)capable they are of managing their symptoms independently without support from clinicians and healthcare services<br>- Make future solutions more inclusive for patients of different ages, literacy levels, mental and physical health conditions.<br>- Include ways to manage the psychological impact of diagnosis and the indirect effects on quality of life<br>- Acknowledge the wider context of the individual beyond their symptoms to encourage a holistic approach to diagnosing CVD.<br>- Reflect on the wide variety of typical and non-typical symptoms related to CVD and provide better information for patients to help improve their ability to recognise when it is appropriate to reach out to health services. |

### Strengths and Limitations

A strength of the present study is the inclusion of both patient and clinician experiences, allowing for a more complete understanding of the barriers preventing accurate and efficient CVD diagnosis. Most existing studies assessed these two groups separately (40,47–50) limiting opportunity to see how needs and requirements compare. A decentralised recruitment strategy also meant the participant sample included a range of individuals from across the country, with a variety of CVD diagnoses (51). The contribution of PPI groups to the design, recruitment, and analysis process, and cross validation of preliminary findings with clinicians at the British Cardiovascular Society Conference increase the transferability of our findings.

Our sample shows a lack of ethnic diversity, particularly in our patient group. This may explain why we did not find patient data to support the role of ‘Cultural Influence’ on patient-clinician interactions. We attempted to remedy this limitation by consulting with a Race and Ethnicity advisory group, who indicated that we might be missing data on culturally specific patient experiences related to family and religion amongst ethnic minorities. Stratified sampling may facilitate adequate ethnic diversity and representation in future research studies.

The use of online recruitment platforms and snowball convenience sampling to recruit our participants may have produced a biased sample of individuals who are more involved in their own healthcare and new technological developments in cardiovascular area. Therefore, our sample may be less representative of patient and clinician populations who are facing challenges in receiving or delivering an accurate and efficient CVD diagnosis. Future research should consider ways to include more seldom heard groups in research investigating contributors to delayed and inaccurate diagnosis of CVD.

There were emerging trends of gender-specific experiences from our patient group that require further investigation, especially given the growing literature exposing how women are at a greater disadvantage when it comes to receiving an accurate and timely CVD diagnosis (48,50,52,53).

## CONCLUSIONS

Several considerations have been suggested for future technological developments striving to address existing contributors to challenges patients and clinicians face to improve the accuracy and efficiency of CVD diagnosis. This study provides insight into patient and clinician experiences of these challenges, and further is required to enhance our understanding of how these experiences differ between ethnic groups and genders.

## Data Availability

Data will be made publicly available upon completion of the primary research aims.

## ACKNOWLEDGEMENTS

We would like to thank the two patient and public involvement groups that helped to inform the design of this study: The NIHR Maudsley Biomedical Research Centre’s Race, Ethnicity and Diversity (READ) advisory group and the Sheffield-based Cardiology Patient group. We would also like to thank Dr Valerie de Angel for sharing her R code to help us create Figure 2 (see Appendix 4) and to thank Helen Denney and Amber Ford for convening the Sheffield patient group and for administrative assistance.

## AUTHOR STATEMENT

### Conceptualisation

F Matcham, TJ Chico, J Condell, O Buckley, V Diaz, R Van Arkel

### Methodology

F Matcham, TJ Chico

### Investigation

K Abdullayev, M Manktelow, O Gorvett, A Sochiera, L Laidlaw

### Writing – original draft

K Abdullayev

### Writing – review and editing

F Matcham, T.J Chico, L Laidlaw, O Gorvett, A Sochiera, M Manktelow, J Condell, O Buckley, V Diaz, R Van Arkel

### Supervision

F Matcham, TJ Chico

### Project Administration

K Abdullayev, F Matcham Funding Acqusition: F Matcham, TJ Chico, J Condell, O Buckley, V Diaz, R Van Arkel

## FUNDING STATEMENT

This work is supported by the UK Engineering and Physical Sciences Research Council (EPSRC) (grant number: EP/X000257/1).

## COMPETING INTERESTS STATEMENT

There are no conflicts of interest.

## DATA SHARING STATEMENT

Data will be made available on request.

## Strengths and Limitations

- The inclusion of both patients and clinicians in our study population is a strength of our study as it allows a more complete understanding of the barriers preventing accurate and efficient CVD diagnosis and comparison between the findings from these groups.
- The use of a decentralised recruitment strategy meant the participant sample included a range of individuals from across the UK with a variety of different CVD and professional experiences.
- The use of online recruitment platforms and snowball convenience sampling to recruit our participants may have produced a biased sample of individuals who are more involved in their own healthcare and new technological developments in cardiovascular area.
- The lack of appropriate recruitment methods to ensure ethnic diversity within our patient sample means our sample may not be representative of culturally specific patient experiences, despite our attempt to remedy this by consulting with a Race and Ethnicity advisory group.

## Notes

### Competing Interest Statement

The authors have declared no competing interest.

