## Appendix 3 for "Stakeholder perspectives on contributors to delayed and inaccurate diagnosis of cardiovascular disease: a UK-based qualitative study"

| **Theme 1: Symptom Interpretation** | | | |
| --- | --- | --- | --- |
| **Subthemes** | **Description** | **References** | |
|  |  | **Patient Experience** | **Clinician Experience** |
| Accuracy of Patient Information | Obtaining accurate patient data is a challenge for clinicians, and accurately communicating and interpreting their symptoms is a concern for patients. False patient information can result in delayed or inaccurate diagnosis and poorer patient experience. | *“But yeah, I find it so difficult, and it really concerns me then, when I'm sat in front of the person who needs to know what has been happening that I can't articulate it very accurately.”*  *“I purchased myself like one of these blood pressure monitors for at home, and I used to take it almost daily, and the readings were within normal bounds. But whenever I went to the doctor’s it was never under 160 over 92 or something and I discovered white coat syndrome at that juncture and I pointed it out to him. So one of the nurses, the the nurse practitioner she tried me on both the the blood pressure monitor, the electric one and on the hand held one. And whenever she did it on the hand held one, it was about 25 points lower and she did it within within 10 minutes.”*  *“I didn't have any perceived symptoms that I knew about although a couple of weeks later my secretary did say you did try and get a car park close to work because you didn't, you got breathless walking to work”*  *“When I got the recording I found that I had said a lot of things I didn't remember saying and I remembered saying things I haven't said at all.”*  *“In the interim I didn't think I had any symptoms but it turned out my blood pressure was higher than it probably should have been. I hadn't picked up these symptoms. I haven't picked up the fact that you know, I didn't go to the doctor and say I think my blood pressure is higher than it should be, because I didn't know I haven't noticed that it was, so I I think that's where something might have been picked up that I wasn't aware of for me.”* | *“I think probably often it's getting an accurate history from the patient as to what's happening, so you can I guess you kind of imagine with like Covid and things, so people are saying, I feel breathless. And you, you kind of having to pick out, is this a respiratory problem? Is it, you know, is it a heart problem? What's going on is this, you know, because lots of people are kind of breathless and getting chest pains after Covid. And so you're trying to kind of pick out from that, is it, have you got some post viral problems? Is it underlying heart disease? Because Covid can make that worse. Is it that people have been quite de-conditioned during the lockdown, like some people have been quite active, but some people have really not done very much for two years, and are now doing a bit more, and like a lot of letters, you get back, it's like oh, I think this person is just breathless because they're deconditioned. It's not actually respiratory or heart disease. So I think it's kind of trying to pick out from the history what's important and what's going on and that's probably the hardest thing”*  *“So let's say you have palpitation, and palpitations happened three times in a year, how on earth, despite your youth, are you going to remember the exact time of onset, time of offset, of the episode of palpitation, and how and how and it affected you at the time when I suddenly be asking you about it next September? I mean so take, I'm taking a lady in her twenties as an example, but the majority of those people will be 85 year olds. So you know my chances of picking up a true reflection of what's going on is much lower.”*  *“Now people are very bad at telling us what distance is so you might ask them what distance, they have no idea what that is. Like you know 200, they might say 200 yards, but you know who uses yards actually, but actually it could be anything”*  *“We find that's quite inaccurate in terms of people's estimate.”*  *“Maybe cultural, personal, whatever it is so sometimes, on some occasions people might um uh not give you all the information. They might choose not to. They might not, you know, and and it might not affect the diagnosis as such, but it might affect how you might go about discussing or agreeing to a treatment plan. So if somebody thinks that if they tell me they have certain symptoms, I'm more likely to go down a certain route or not then you know, that obviously affects it but it may not affect the actual diagnosis because let’s say, we realise that but it obviously affects the conversation.“*  *“Well, obviously, oh, for culturally, alcohol is a massive issue in our cohort of patients. The natural will to reduce down the amount that they’re saying that they're actually drinking, the concept in [City] of what is normal drinking is not normal.”*  *“Try and make sure I do a decent vape history nowadays because people will tell you they don't smoke, but they're vaping. That's a big area where we don't really know where we're at with all of that.”*  *“Therefore, they they they don't want to have a diagnosis of ischaemic heart disease, despite the fact, it's looking at them straight in the face. Sometimes ignorance on on behalf of the the the patients as well, although I’m saying a lot of them know what they they have, there's other patients will, will, will not, or will choose not to to know what what's going on, and so like lack of knowledge as well.”* |
| Ambiguity of Symptoms and Misdiagnosis | Patients and clinicians both describe the ambiguity of potential heart disease symptoms and how this can cause confusion and increase the likelihood of misdiagnosing the condition, which is a common experience amongst patients. | *“I got to the doctors I was the last patient that he saw that evening, so he made extra space for me and unfortunately, he misdiagnosed it. He thought it was a just severe indigestion and he gave me something for that.”*  *“I've never had any sort of problem other than with the nitro-glycerine tubes they give you. You're told that if you feel chest pain coming on, you should give yourself a couple of sprays under your tongue and the pain will disappear. About four months after the event, I was walking along the road when I thought I was having chest pains. I did what was instructed and gave myself the spray and had the worst headache I've ever had in my life, which apparently is what happens to you if you're not having a heart attack and you use the the spray, it gives you a boomer of a headache.”*  *“I’d had one or two little occasions in between then, from when I'd been hospital in hospital to the GP, when I thought, hmm was that the same thing again but they only lasted very short period of time.”*  *“I find they were quite subtle, and it's easy, you know, have I I got one at the moment? and then I’m there fingers on me. I can't find my bloody pulse!”*  *“The the main thing that concerns me was the manner in which it happened. At the time of the incident I was sitting. I wasn't in an excited state. I hadn’t been doing any form of physical activity before and I've often wondered, I used to suffer from indigestion and I I I sometimes wonder if what I put down to indigestion might have been something else because I still find it strange to this day that you can be sitting doing nothing and, bam! You have in effect, a heart attack. It's just it it it makes me think, and I'm always conscious of it…But it's a concern. It's the uncertainty of what really happened because it doesn't quite get explained to you.”*  *“I look back too and believe now I had episodes before I finally sought help”*  *“But I used that for a long time because you can think can I feel this, or is it because my heart is beating a bit quicker because I’m stressed? And is this AF or am I just getting myself worked up with it? So you kind of doubt your reliability, don't you?”*  *“Then I was on medication and it was that, shall I go to hospital? Shall I not?”*  *“I found myself leaving things at the bottom of the stairs so that I was only going up and down once rather than going up and down and up and down. The only symptom that I had was, I had burning in my throat… when I did any kind of exercise. So if I climbed the stairs I would have burning in my throat. But no other pain no pain anywhere. Nothing like that at all...Eventually went to my doctor, after I’d planted three fruit trees in the garden at the weekend in heavy clay soil and discovered I could only do a bit, and I felt really tired, so took me the whole weekend to do it. How crazy was I even doing it? But obviously I didn't know what was happening.”*  *“Now I used to be a nurse many years ago, learning disability nurse, and had been a social worker, but I’d never come across anybody talking, I'd work with a lot of people with heart attacks, never came across anybody talking about the symptom of burning in their throat. So it it took a long time, really, for it to occur to me that that was what was happening.”*  *“I woke up on Good Friday morning with a what I thought was obviously it was a punctured lung from playing football.”*  *“The first real problem that I had was that in 2015 having three years earlier thought I might have had a problem with my heart and being dismissed, I had a stroke and spent 10 days in the stroke ward of the local hospital”*  *“I was referred just to a general consultant, who thought that I actually had cirrhosis of the liver…I had a liver biopsy and I didn't have cirrhosis to the liver…it did turn out to be a heart condition”*  *“Since then, I've been okay-ish. I still get the palpitations, but no one really knows why do I get them. I can't work out myself either.”*  *“Prior to that I was just doing my normal routine, bicycle and things like that, riding the bike. And then suddenly, I couldn't couldn’t understand why I was getting more more more breathless.”* | *“You kind of imagine with like Covid and things, so people are saying, I feel breathless. And you, you kind of having to pick out, is this a respiratory problem? Is it, you know, is it a heart problem? What's going on is this, you know, because lots of people are kind of breathless and getting chest pains after Covid. And so you're trying to kind of pick out from that, is it, have you got some post viral problems? Is it underlying heart disease? Because Covid can make that worse. Is it that people have been quite de-conditioned during the lockdown, like some people have been quite active, but some people have really not done very much for teo years, and are now doing a bit more, and like a lot of letters, you get back, it's like oh, I think this person is just breathless because they're deconditioned. It's not actually respiratory or heart disease.”*  *“So, for example, my man last week that was, he rang the practice, he was deaf, and he came into the practice, and his ECG was normal but the but the history was just barn door ischemic heart disease.”*  *“I use a recent example with a, was a patient who was complaining of neck pain, and when he was playing golf, and thought thought it was an arthritis thing, so it it could be a a general symptom.”*  *“A lot of cardiovascular conditions exist and and patients know nothing about them they just, you know, like hypertension, for example. So how do we get those patients to come to see us to to get this investigation fitted to you know, are we should we be proactively looking for them.”* |
| Clinician Dismissal of Symptoms | Patients expressed feelings of being dismissed by clinical staff when their symptoms didn’t match the status quo, which led to delayed access to the appropriate tests or treatment and increased anxiety and frustration. | *“I don't think I have ever bought up any concerns with my doctor about my heart. I thought I was, but they took very little notice of it. And in fact, the first real problem that I had was that in 2015 having three years earlier thought I might have had a problem with my heart and being dismissed, I had a stroke and spent 10 days in the stroke ward of the local hospital”*  *“I was having constant migraine all the time no one did anything, not even the doctors, until it was so bad, and then they realized this migraine wasn't migraine, it was actually blood pressure and heart disease, I was having a heart attack. That was my initial start of it.”*  *“Just woke up one night and just knew something wasn't right and when the ambulance came and they said, what's wrong? And I said, I don't know but something is and they went oh, you'll be all right.”*  *“One of the consultants told me that I don't look like a heart problem because I am still young, and I don't, I'm not overweight, or I don't drink or alcohol anything like that. So I shouldn't have a heart problem basically. So I was like, oh, okay, so they discharged me two years ago.”*  *“Then when I explained, they said, they didn't take it seriously “oh okay” well they eventually they came, "we'll take you to hospital, then we'll find out from there”. So when they did blood test then it showed that I had I had had heart attack and troponin level was 114.”*  *“I had no pain in my chest or in my arm or anything and they didn't believe that anything was wrong until they had done the blood test and it showed that I had a heart attack.”*  *“I walked to the ambulance fine, but the ambulance just didn't believe that anything was wrong.”*  *“I felt it wasn't taken very seriously.”* | *NA* |
| Diagnostic Overshadowing | Both patients and clinicians acknowledge the role of existing comorbidities in complicating heart disease diagnosis, as well as difficulties with heart disease overshadowing other potential conditions that manifest in addition to the heart disease. The overlap in symptomology acts as a barrier for clinicians, making it difficult to distinguish potential causes and prescribe appropriate treatment. | *“I've often wondered, I used to suffer from indigestion and I I I sometimes wonder if what I put down to indigestion might have been something else”*  *“The big problem I've had is that whenever I I went to my GP and said, I’m not feeling well, he would say, well, you have a heart problem but I'm pretty sure that the last two years I've been suffering from something other than my heart, or in addition to my heart.”*  *“I was diagnosed also with type two diabetes in 2012. So that was three years before I had the first lot of stenting done. Clearly that's a factor, because obviously it makes your blood sticky, and you know none of that helps. I’ve got atherosclerosis which isn't curable, of course, I mean statins and stuff obviously help with that. But you know it's never going to go away. It's not something that goes away.”*  *“I always wanted to get to the root of I always thought arrythmia was related to the anxiety which in theory it is, or it isn't, I don't it's an aspect of it.”*  *“I have a myriad of other problems, and nobody seems interested in nothing.”*  *“I think what one of those dangers is that you, when you've been diagnosed with a heart condition and you you do start to look after your health, when you get, when you do talk to a doctor they will immediately you know, I think the the old saying that you know when when your only tool is a hammer, you know every problem looks like a nail, and it's an it’s it it I my pulmonary embolism, it actually went on, I think I had more than one and but it but they were so focused on the fact that I you got a heart condition that it masked any genuine investigation into what else was wrong with me.”*  *“I think that you know this one that you can look after your health but other things may go wrong with you, and that you may actually find that that people's preoccupation with the fact that your you know you've got a heart condition that is potentially going to mask that because there's an automatic assumption that that's what you're actually suffering from.”*  *“That's something that that from how he explained that happened there is very much lacking in in the NHS, because they focused only on my Type One diabetes, tried to look after that but weren't really concerned with all of the, what I now know, are very considerable and frequent side effects and really that's why they have to spend a lot of money when it all goes wrong.”* | *“You kind of imagine with like Covid and things, so people are saying, I feel breathless. And you, you kind of having to pick out, is this a respiratory problem? Is it, you know, is it a heart problem? What's going on is this, you know, because lots of people are kind of breathless and getting chest pains after Covid. And so you're trying to kind of pick out from that, is it, have you got some post viral problems? Is it underlying heart disease? Because Covid can make that worse. Is it that people have been quite de-conditioned during the lockdown, like some people have been quite active, but some people have really not done very much for teo years, and are now doing a bit more, and like a lot of letters, you get back, it's like oh, I think this person is just breathless because they're deconditioned. It's not actually respiratory or heart disease.”*  *“I I think obviously, there's you know, the chest cardiac symptoms can sometimes be very vague and overlap with other diagnosis. So you've always got to keep an open mind, even if you're you know you've got someone with chest pain, and you've got raised troponin, and you know this could be other causes that are still there as well.”*  *“All these systems can be affected by the heart, and equally the heart can be affecting, affected by them and their illnesses. We call those comorbidities. So is it important to find out more about the remainder? Absolutely. Yes. Absolutely yes, because that helps you all the way, in terms of knowing which diagnoses is the most likely, most common. You also need that information to help you decide what is the most appropriate of a huge battery of tests and a huge battery of interventions”* |
| Experience-based Clinical Intuition | Clinicians often describe how their personal clinical experience guides their decision-making processes during diagnosis and how they respond to out of the ordinary symptom presentation. This highlights a reliance on subjective intuition when guidelines fail to provide adequate support. | *NA* | *“Sometimes, in fact, not infrequently, you may have the diagnosis firmly in your mind but because there are other possibilities, you want to have confirmatory diagnosis by testing, and the tests vary.”*  *“The repetitive bit is that you are taking a history in and in a way that may sound to the outsider as haphazard but it actually is not because it has a basis in terms of our knowledge of the biology, physiology, pharmacology, pathology, and histopathology, and as well as clinical knowledge.”*  *“You examine the patient to confirm your suspicion and confirm it further by further tests to reach a diagnosis, does that answer your question.”*  *“I I think the most in in in medicine in my opinion, seeing the patient, asking the patient the questions about their symptoms, the severity of their symptoms, that would give me a very good idea of whether the patient is getting better, or the same or getting worse.”*  *“Usually, we start from subjective subjective cues first. For instance, again, is going to about going back to a clinical history asking the patient, asking the patient some questions, examining them, and it's quite well accepted that you know in about 80, you know, quite quite often, you know, you may see in a textbook that 80% of the diagnosis in medicine can be made by just taking a good history.”*  *“I think, even in this this day a clinical assessment is really important, and then we may or an examination which tends to give us pretty much a good idea of the diagnosis, and then they we send them on for investigations.”*  *“How do I feel? Quite good? Well generally it's um it's I think it comes with the experience. So now, having done it for a long time, you kind of can gauge better of how people are taking in things, I think and recognising that everybody is very different. So how I might have a conversation with one person or one family might be quite different, based on how you're reading their feedback to it, and I think I found that easier as time goes with experience, I think.”*  *“Then the key thing is the history. Speaking to the patient. Letting the patient often for firstly, for an open question let the patient speak for themselves, describe any symptoms, try not to interrupt them during this period and from there you sort of go from the open questions down to some more sort of closed questions. So, for instance, of chest pain, you know, when when did it start? What's it like? Has it spread anywhere? Anything make it better or anything make it worse? Any associated symptoms?”*  *“So, I think the you know the the patient's opinion I think, is first of all, it's most important. What they feel is going on. What they're worried about. These things are, you know, so it's an important to involve the patient at any point.”*  *“We got access to proBNP quite early, and I've been using that as a, as a diagnostic tool, to sort of try to assist me, to get the good insight into the role of proBNP and I know the role of, how proBNP can be contaminated by things like atrial fibrillation or renal disease, so I’ll try and take a history, ECG, lipids”*  *“I’m a big fan of 24hour blood pressure monitors. We were well ahead of the curve and early adopters to the concept of 24 hour blood pressure monitoring. And again, because I had a background in diabetes, and they were very fond of 24 hour blood pressure monitors, just to have a look at autonomic tone. I was really kind of, came to the practice about ten years ago, and I said, folks, I think we need to be doing 24 hour blood pressure monitors, not treating people based on one-off readings”*  *“I’m a big fan of 24 hour blood pressure monitoring, both to assess for daytime readings, for night time readings and their nocturnal drive. So for me I’m a big fan of blood pressure.”*  *“if I’m looking at cardiovascular disease, for me eGFR is a massive tool, underrated and ignored a fair bit I think, but as somebody's eGFR, relatively young, is going off, that, to me is a cardiovascular cancer waiting to happen.”*  *“I see upwards of 30 people a day I I can normally detect within seconds where to pitch something. You know it's a skill that you learn in primary care, in my opinion.”*  *“I tend to, you know I diarize those kinds of people, so I keep a kind of close eye on it and and understand that they're not they're not, what I've learned from doing this now for 25 years is it’s very it's it's not very likely that somebody with a cardiac condition or cardiac disease is sitting at home thinking I need to get all this sorted and I just can't be bothered, there's myriad reasons why they don't engage.”*  *“I will put whatever investigations that I feel are needed into train and we’ll sort out further investigations and referrals from there.”*  *“Heart disease diagnosis from my point of view, is normally made on on history”*  *“Then you have the getting into the hospital investigation or secondary care investigation obstacles too, they they follow quite strict guidelines, which I find a little bit of an obstacle because as a clinician, I may well feel very strongly that this is a ischaemic heart disease and and you make that case in your in your referral, but if it doesn't meet the criteria, and then it is rejected.”* |
| Gender and Age Overshadowing | Patients expressed frustration with how their gender or age resulted in false assumptions about their cardiovascular health from clinicians, but also how their age affected their personal ability to distinguish between natural regression of physical health or a symptom of heart disease. Meanwhile clinicians describe how they tailor their approach to diagnosis based on demographic factors, suggesting this is an important factor in their decision-making process when diagnosing a patient with a heart disease. | *“For 20 odd years, I never thought about it but of late the last couple of years, I suppose as you get slightly older, you do start to think well maybe I should, maybe I shouldn't, it's I feel like I'm at a cross roads on occasion, you know. Should I phone the doctor, should I not?”*  *“As as you get older, the the probability that you will increase in ailments goes up.”*  *“I'm struggling now at 76, I’m struggling to walk up steep hills. But at 76 that's probably expected, anyway.”*  *“You sort of just accept that you're getting older, slowing down and it's nothing to do with the heart. It it's just general slowing down with the body, and the fear you could be suffering from heart disease.”*  *“Physically it's it's impossible, because, you're as you get older, you deteriorate even more in health, and with the added problems of heart disease, it it becomes a major problem with such as breathlessness”*  *“So, I gave up on NHS to be honest. I thought it was only me. One of the consultants told me that I don't look like a heart problem because I am still young, and I don't, I'm not overweight, or I don't drink or alcohol anything like that. So I shouldn't have a heart problem basically. So I was like, oh, okay, so they discharged me two years ago.”*  *“I suspect men are worse, they don’t go to see doctors. Don't like them. Oh, no, they might like the doctor, but they don't like what the doctors might tell them.”*  *“Also the lack of symptoms! If you haven't got symptoms of course you can't really, or or maybe there's not enough education about the kind of more obscure, because apparently, it’s quite common for women in particular not to have the kind of standard you know, pain down the arm and that kind of stuff. So maybe there's not enough knowledge, not enough information given, general information given you know, to the wider public about the the possible symptoms of heart attack that it is not just, you get all the traditional ones about the pain in the chest, the pain down the arm, the pain in the back. But I haven't heard anybody, even with my experience, talking like I said, about having pain in your throat, pain in the jaw, but not not kind of, but then I was told by the cardiologist, that's very common in women So I think why isn't that, why isn’t that more widely known? So I suppose that's the thing.”*  *“There seems to be a stereotype of what is a heart attack, and the symptoms that the doctors are seem to be very stereotyped and focused on what the symptoms are of a heart attack for both men and women and yet I never experienced those symptoms that’s in the book that these doctors learn from. A woman's heart attack is both, on all three of my occasions have been different to what the book and the doctor states.”* | *“Very organised man in his early eighties but when you ask the questions further it turns out that this person's life has become confined to his home not through physical disability but because of frailty. A frailty is a very wide diagnosis, but it encompasses ailments, and illnesses, and deficiencies in more than one place and that would make the general appearance, as well as the behaviour of the said patient, as frail.”*  *“You know if you've got a diabetic, smoker, then that's you know more likely to be down one diagnosis than another, and their age and frailty and then also, there's a premorbid status as well, like currently in [City] so we get a lot of very elderly patients who are very stoic but actually I've got quite a lot of comorbidities.”* |
| Holistic Considerations | Patients had different experiences when it came to clinicians taking into account several factors related to their health, with some feeling like clinicians only cared about specific numbers and tests, while others felt that their mental health was factored in to their care.  Clinicians also discussed holistic approaches to diagnosis to varying degrees, with some highlighting the importance of considering patient mental health and quality of life, and others focusing almost exclusively on results of investigations and physical presentation of symptoms. | *“The NHS provided six sessions with a psychologist, which was all right. It was, it was pretty good, but that was that was all that they could give me. So I then went privately and I I actually found that I did need that that support from, it was really talking therapies, really that's uhhh what I needed. But it was important. It was, looking back, quite an important factor in managing the condition up until I had the ablations. Once I’d had the ablations, everything was a whole lot lot better but prior to that, when I was still having symptoms on a more regular basis, it was very troublesome and, and I did need that support.”*  *“My sleep was so disrupted”*  *“I always wanted to get to the root of*  *I always thought arrythmia was related to the anxiety which in theory it is, or it isn't, I don't it's an aspect of it.”*  *“It's never really it's never really tied in with the mental health and and I think it should be, since I work in mental health”*  *“Certainly at [Hospital-name] that’s very much recognized, because that was talked about a lot for me the few days I was in about how I was feeling emotionally, and stuff like that. I was given opportunity to talk about how I was feeling emotionally.”*  *“Their job seems to be more physical in terms of, you know, treating the condition rather than, you know, the mental aspect of it or the ongoing aspects of it”*  *“It would be good if there was suitable after care to deal with the mental health aspect”* | *“I think it's kind of trying to pick out from the history what's important and what's going on and that's probably the hardest thing”*  *“The patient turns out to be a smoker and two members of his family have had illnesses that he was not sure of that led to their premature death and you examine the patient and start eliciting signs of congestion on the lungs”*  *“The repetitive bit is that you are taking a history in and in a way that may sound to the outsider as haphazard but it actually is not because it has a basis in terms of our knowledge of the biology, physiology, pharmacology, pathology, and histopathology, and as well as clinical knowledge”*  *“Very organised man in his early eighties but when you ask the questions further it turns out that this person's life has become confined to his home not through physical disability but because of frailty. A frailty is a very wide diagnosis, but it encompasses ailments, and illnesses, and deficiencies in more than one place and that would make the general appearance, as well as the behaviour of the said patient, as frail.”*  *“It's also about their how the how the symptoms and the condition affects their life, affects their daily activities. This is important to measure quality of life… this the other thing that is getting more important more and more important is this, is patient satisfaction. So this this this addition of tools compared to the conventional medicine that we just asked, how bad is the pain? How bad is the condition? Yeah. So so depending on you know how how they affect the quality of life and how satisfied the patient is.”*  *“You know their general wellbeing, their life, and how the condition is going to affect them. And then, once I've got a diagnosis, I'll try to tailor, you know I do my best to tailor my treatment plan according to their expectation, according to their background. You know, to get to get the best out of the the treatment for the for the individual patient.”*  *“But of course that is in the setting of the laboratory so when you walk outside a lot of things vary right? Whether you're walking up the slope, down a slope. Is it windy? Is it cold that day? Are they carrying stuff? And, at the end of the day those are the things that are relevant because it's related to what they need to do in their daily lives.”*  *“We don't routinely do quality of life questionnaires in the clinic, partly because of time, but also there are problems using some of these tools as well. ”*  *“Well, it's key to know what the other comorbidities are. So it's very different seeing a 40 year old with no comorbidities versus a 96 year old, who's in a care home with lots of comorbidities, including dementia and perhaps cancer diagnosis or lung disease. So it, this sort of context you know the the patient specific context of their cardiac disease is crucial to knowing what the next stage is, and indeed, whether we need to see them or not.”*  *“I think the you know the the patient's opinion I think, is first of all, it's most important. What they feel is going on. What they're worried about. These things are, you know, so it's an important to involve the patient at any point. From there, I also want to look at the patient’s past medical history, their comorbidities, you know, their lifestyle choices. These things can often influence what's their diagnosis.”*  *“At that time, I tend to kind of do a more, a more intense assessment of you know, what the family, you know there might be missing data. You know what's the family history, being really specific. Is it really a premature history? Is it on, you know, I said, a woman under 60, is it a man under 50 and get all that data on board, just to to make sure that the Q-risk is accurate”*  *“When you spend five minutes interrogating them, they’ve just got a a death or a grief or something that’s happened.”*  *“So, you're you're looking at patient background, and their socio-economic background, their personal family history, as reg-, as as regards to ischaemic heart disease and and the the story that they're telling you, so you're talking about smoking as well, diet, stress, exercise, basically the the the whole picture.”*  *“I suppose they're you're you're assessing the patient, you know you're looking at the big, the whole picture rather than just the set of numbers and and things in isolation so you know you you're you're looking at the risk factors the patient might have, you know, such as maybe the family history or their lifestyle, or you know, I suppose that includes their diet or the level of exercise, or if they smoke, or and maybe they have a sort of background of other health problems hypertension, possibly and hyperlipidaemia and so I mean, this is sort of you have all that information I mean I work as a GP so we have a lot of that information without having to repeatedly ask the patient so yeah, I mean you, you you're gathering all that together in your decision as well.”* |
| Incomplete Patient Information | Both patients and clinicians expressed concerns with having sufficient data to inform an accurate diagnosis. For patients, this involved worries surrounding the way their data was collected and whether it was enough to inform the diagnosis. For clinicians, there was a concern that patients were either not able, or did not want, to give their doctor a complete history which impeded on their ability to make an informed decision on their condition. | *“I just remember at the time being surprised how inexact the way the diagnosed was, that they were relying on a 24 hour period of time to decide what it what it was, or whatever. If I didn't have an episode, there would be nothing, nothing to gather, and, as it happened, there was barely a blip in the 24- or 48-hour period, but they still diagnosed paroxysmal atrial fibrillation.”*  *“Then the cardiologist would be asking, and I'd be thinking, blimey. I'm only giving him half an information here, how is he going to help me when I can't be exact?”* | *“And the the area I work is not the most affluent so often, it's they, they don't even know, even them trying to say to them, you need to go and ask your family about your family history. And do, you know, go and ask whether anyone's got high blood pressure? Have they got diabetes, are your family on statins? That kind of question, just to get a bit more of a a kind of a picture of what's actually happening in their family.”*  *“Maybe cultural, personal, whatever it is so sometimes, on some occasions people might um uh not give you all the information. They might choose not to. They might not, you know, and and it might not affect the diagnosis as such, but it might affect how you might go about discussing or agreeing to a treatment plan. So if somebody thinks that if they tell me they have certain symptoms, I'm more likely to go down a certain route or not then you know, that obviously affects it but it may not affect the actual diagnosis because  let's say, we realise that but it obviously affects the conversation.”*  *“I think we need to be doing 24 hour blood pressure monitors, not treating people based on one-off readings”*  *“Big problem for doctors as patients talk in terms of, and obviously again with very poor and poverty ridden demographic, a lot of them roll their own cigarettes, and they say I only take half an ounce, I don't even know what that means. I'm like well, what is that, you know? So I will ask the patient to tell me what that would be roughly in terms of normal cigarettes.”*  *“I've had about I can easily say about five patients that were admitted to a surg-, a medical ward and started on blood pressure medication based on blood pressures that don’t even make NICE guidelines and I bring them, they come out to me, and I’m like I don't - can we just put them on a bloody monitor, and then itt's normal. And when you spend five minutes interrogating them, they’ve just got a a death or a grief or something that’s happened.”* |
| Patient Intuition | Some patient stories showed that reliance on personal intuition played a crucial role in patient interpretation of their symptoms. This highlights the subjectivity of symptom interpretation and potential difficulties associated with communicating these feelings to clinicians in a medical setting. | *“Called up the local surgery and said, could I come and see the doctor? Because at the time I didn't recognise it as a heart attack, because I never thought that would happen to me.”*  *“But then I kept going, saying I was feeling unwell and sweating, and I didn't think for one minute I'd had a heart attack, but I was diagnosed with AF, paroxysmal AF.”*  *“I look back too and believe now I had episodes before I finally sought help”*  *“a couple of weeks later my secretary did say you did try and get a car park close to work because you didn't, you got breathless walking to work. Basically I didn't have any symptoms that I I perceived.”*  *“I was 100% sure, I was having a heart attack, you know, when somebody’s sitting on your chest, you know you're having a heart attack. You know it’s not, there’s no question about it, as far as I was concerned.”*  *“I think there is probably the main one, you don't, you sort of just accept that you're getting older, slowing down and it's nothing to do with the heart. It it's just general slowing down with the body, and the fear you could be suffering from heart disease. I suspect men are worse, they don’t go to see doctors. Don't like them. Oh, no, they might like the doctor, but they don't like what the doctors might tell them.”*  *“Just woke up one night and just knew something wasn't right and when the ambulance came and they said, what's wrong? And I said, I don't know but something is.”*  *“Anybody who's been ill, and I trust we were all in that situation, can probably identify with the fact that when you're ill you don't know you're ill. As far as you’re concerned, you’re fully compos mentis and able but you’re not. You're as mad as a March hare.”* | *NA* |
| Variety in Initial Symptoms | Each patient had a unique story of the series of events and symptoms that lead to their heart disease diagnosis and treatment. This highlights the importance of considering the wide range of symptoms that can pre-empt a heart episode or condition. | *“I kept going, saying I was feeling unwell and sweating”*  *“I was just feeling really rubbish, and felt,*  *carried on feeling rubbish till about two or three o'clock and kept feeling my chest and thinking there's something not right.”*  *“I started having events, tachycardia, feeling faint.”*  *“I collapsed in the house with a very, very fast, heart rate, 200 beats per minute.”*  *“ I had burning in my throat here in my throat when I did any kind of exercise. If I climbed the stairs I would have burning in my throat. But no other pain no pain anywhere. Nothing like that at all.”*  *“the pain came in my jaw”*  *“I had swollen legs and very I was very, very breathless.”*  *“I was having constant migraine all the time no one did anything, not even the doctors, until it was so bad, and then they realized this migraine wasn't migraine, it was actually blood pressure and heart disease, I was having a heart attack. That was my initial start of it.”*  *“basically I get palpitations and sometimes followed by heart attacks.”*  *“Mine was like I had the palpitations lasting for five hours and then we were still hoping that it would stop by itself yeah… I could measure with my Fitbit, it it went up to 200 per minute. Yeah. And then I walk then my legs gave up. I couldn't walk any longer, but I was desperate to get to the bathroom I crawled, I crawled, and and then, all of a sudden, cold sweat. I was sweating away, literally soaking wet but cold and shivering at the same time and I wanted to vomit, nausea. And I had a phone with with with me. I wanted to call ambulance. I dialled the number, but I couldn't speak. I just couldn't speak. My speech was slurred I couldn't, whatever I said it didn't make any sense…And second time when it happened very similar symptoms, my legs go first I just collapse.”*  *“I got the sweats. It was like you was under a shower. I was dripping so much. And yeah, I was freezing. But there was no pain. I had no pain in my chest or in my arm or anything and they didn't believe that anything was wrong until they had done the blood test and it showed that I had a heart attack and it was totally out of the blue. I just woke up in the middle of the night. But yeah, just there's no pain”*  *“All my symptoms were hidden. The only way I knew I was actually having a heart attack was severe acid reflux.”*  *“I had night sweats and chest tightness for a number of months before my heart attack. Then, on the day I had really heavy sweating back and shoulder pain and episodes of chest pain.”*  *“I've got an erratic heartbeat so what's called at Atrial Fibrillation. The only time I knew about it was when I went for my annual check-up at the doctor and the doctor sort of listened to my heart, and said, oh, you've got a funny heartbeat.”*  *“I I was getting a little bit breathless. Prior to that I was just doing my normal routine, bicycle and things like that, riding the bike. And then suddenly, I couldn't couldn’t understand why I was getting more more more breathless.”*  *“those were the symptoms breathlessness or lack of not breathing, being able to breathe so easily on exertion. and it was picked up from that.”* | *NA* |
| CL Clinician. P Patient. Redacted text shown in []. | | | |

| **Theme 2: Patient Characteristics** | | | |
| --- | --- | --- | --- |
| **Subthemes** | **Description** | **References** | |
|  |  | **Patient Experience** | **Clinician Experience** |
| Anxieties and Fears | Many patients describe the fear and anxiety they had to deal with in the period around the diagnosis of their heart condition and how this impacted their wellbeing. Meanwhile, clinicians also acknowledged patient anxiety as an important factor to consider when communicating a diagnosis and managing patient emotions and expectations. The data suggests that failure to consider the emotional impact of heart diseases on patient wellbeing will result in worse patient outcomes. | *“It didn't feel like that at the start. It was probably quite terrifying. I had 3 boys under the age of 10 at the time”*  *“Since I got the defibrillator implanted the first 6 months, maybe a year I was particularly conscious of any exercise or any, anything that could raise the heart rate and could result in the defibrillator giving me a shock. So I was definitely monitoring heart rate, pulse, consciously, subconsciously, walking up a flight of stairs, going for ride on my bike, every day was a little bit of a how much can I do? What's possible? What's not possible? What's going to trigger it? What isn't?”*  *“It's just questions you start to ask yourself, it it raises doubts in your own mind, and I think that's the worst thing”*  *“A lot of anxiety around the condition, particularly whilst or once I was diagnosed”*  *“Or I don't feel great, but can't go to that party, because I just don't feel as though I want to mix. So I felt I was fighting anxiety that was getting really quite troublesome.”*  *“For the first 48-72 hours after your incidence, to me the fear was crippling. Because you don't understand what's happened, you’re to an extent terrified to do anything that may invoke another incident”*  *“But you're hooked up to all sorts of monitors and when a visitor would come in, I’d say, Hi! and immediately one of the nurses would say calm down, calm down, your heart rate’s gone up, and the impact that actually had on you, is it's just, I found the fear was the worst thing for 72, 48 to 72 hours.”*  *“I had no warning before, absolutely no warning before, that it was coming. And that was the worry for me. I never really worried about it,”*  *“It stopped me to a halt, really because I was so scared.”*  *“Basically everybody in the hospital tells you what they're going to do and each and every one of them puts the fear of God in you”*  *“You go through everything else like heart attacks, and thrombosis and it's sort of it's quite a scary thing, to, an operation to go through. I think that the professionals don't quite realize how much they're frightening the patients before the operation. I think that was my biggest concern.”*  *“But the fear of being in the house on my own knowing, you know, I was 100% sure, I was having a heart attack, you know, when somebody’s sitting on your chest, you know you're having a heart attack.”*  *“all you're concerned about is basically you've had an heart attack, you're either going to live or you're going to die. So whatever they do you accept, because you know, the alternative is the end”*  *“I remember when I first got it, was was diagnosed, so I knew what the the conditions were and you go on the Internet and it it's it's really quite scary on the Internet because you read all these horror stories about, you know, I think that right I you know, I read about a number of places that that I'd be, and there was a 40% chance of me being dead within two years.”*  *“I think that that that was the worst bit for me when I was first diagnosed. I didn't just didn't know what the future was going to hold or how long I was going to survive.”*  *“You never know when it will happen again. It's always on your mind after you have had an event, or if the next one will be worse, or will be a worst scenario”* | *“I don't hide things from them but the reason for that is not to frighten them, even though they may well get frightened”*  *“They sometimes are very frightened so, they don't come back to get bloods checked,”*  *“So we're we're quite pragmatic and proactive in trying to reduce that anxiety of those concerns.”*  *“Most patients don't get it, so they get panicky when they see numbers like 90, and I’m not panicking”*  *“I think it's about trying to remove the fear from the from the patient trying to de-escalate them, because you, you'll often see panic, particularly in the part of the world that we live in, where there's a very high incidence of ischaemic heart disease and explain to patients that the important thing is making the diagnosis and then making the changes that are needed to reduce the impact of that diagnosis. So the the the management, to my mind, is all about reducing fear to allow the patient to engage in the management of their of their condition.”*  *“So it's emphasizing the seriousness, but at the same time trying to keep the worry levels down.”*  *“We've talked about fear on on the the patient's behalf, reticence the fact that they don't want to have a diagnosis of ischaemic heart disease.”* |
| Individual Patient Differences | Both patients and clinicians discussed the importance of considering individual patient differences throughout the diagnosis process, from initial appointments to treatment plans. There was a mutual understanding amongst patients and clinicians that a one-for-all approach does not allow for appropriate patient care. | *“I had 3 boys under the age of 10 at the time”*  *“There were lots of really good tips. You just have to adjust them to your lifestyle as an individual.”*  *“As an engineer I much prefer to see things and so being able to see yes, that's what's happening. Look that that is what a normal one looks like! I like to to see the the evidence, the ultrasound scans that they did that prove that actually, physically, your heart was fine. It's it's your electrics that's shot on it. I I find I I do appreciate seeing those things,”*  *“I remember them discussing Warfarin, and why it was important in the chart that you were assessed, if you were a woman, if you had a certain age, you know risk factors.”*  *“I’m on something called “Patient View”, which is again just about to change its name to “Patient Knows Best” I think it's going to be called, so I had access to all my blood results. So, being an ex-nurse, obviously I look at everything”*  *“My greatest challenge was, I live on my own”*  *“I was in a hospital for a week and came out with medication which really upset my life. So began over a period of time weaning me off it”*  *“I was 24 in a hospital, youngest by many years”*  *“obviously in social work, I’m used to dealing with medical professionals a lot, I really valued all the information that I was being given.”*  *“Yeah can I just say that everybody is different. And everyone is totally different and is what they want, what they want to know, how much they want to know, and how they want to know it. I personally would much prefer seeing a doctor rather than listening to a computer telling how poorly I am. Other people would much prefer the computer rather than the doctor. And I just think we need to bear in mind that all human beings are different.”*  *“I act as a carer 24/7 for my wife”* | *“And patients themselves are quite variable. So some if you said there’s things you can do at home would really engage with that. Others would just be. no, no I just want to see a specialist to be told what the problem is.”*  *“I think they're probably more tolerant of certain things in some respects.”*  *“It depends here on the scenario, again and depends on what condition it is.”*  *“When somebody is frail there will be things that you would wish not to offer to. So somebody who's got severe frailty, the last thing you want is to offer them complicated and complex surgery from which a young person might recover in five days, but they may not recover in eighty days. And if they were to have a serious heart condition that is likely to shorten their lives then losing six months is not unimportant”*  *“That shouldn't be applied across the board and not everybody should be deprived from being seen but some can.”*  *“Even though the you know, let's say I'm only, it’s for the same diagnosis, different patients might take it very differently and different patients might react very differently, might be able to, the coping mechanisms will be different. The questions they're going to ask, may be different, and how the condition is going to affect their life will be, you know, will be different, even though it's it's the same condition. It's the same diagnosis. So as a clinician when I first meet the patient I keep my mind very open and try to gauge how you know their background, their expectation. You know their general wellbeing, their life, and how the condition is going to affect them. And then, once I've got a diagnosis, I'll try to tailor, you know I do my best to tailor my treatment plan according to their expectation, according to their background. You know, to get to get the best out of the the treatment for the for the individual patient.”*  *“You need to adjust how this treatment. Is this treatment going to be fit for this patient? Is this something that the patient expects? Is there anything that the patients, they see that this treatment is not is not suitable for them. Is the treatment too high risk for the patient to accept?”*  *“Of course, the problem here is a lot of patients don’t always have their phones with them, so they’re popping out, they might not. So they're not necessary of the age group that have their phones glued to them Haha. So they say, well, you know it's very well, but I've brought my phone today because I'm coming to the hospital for an appointment, some days when I'm just popping out, I won’t.”*  *“Well generally it's um it's I think it comes with the experience. So now, having done it for a long time, you kind of can gauge better of how people are taking in things, I think and recognising that everybody is very different. So how I might have a conversation with one person or one family might be quite different, based on how you're reading their feedback to it, and I think I found that easier as time goes with experience, I think.”*  *“Well, it's key to know what the other comorbidities are. So it's very different seeing a 40 year old with no comorbidities versus a 96 year old, who's in a care home with lots of comorbidities, including dementia and perhaps cancer diagnosis or lung disease. So it, this sort of context you know the the patient specific context of their cardiac disease is crucial to knowing what the next stage is, and indeed, whether we need to see them or not.”*  *“Like every every patient's different, and also depends on every situation”*  *“So in terms of making the diagnosis, you know, you can make a you know, diagnosis of a an MI, and you know and you know theoretically they should go for angiogram but when they're 97 and  you know they have lots of co-morbidities and you know they they don't really want to leave the island than these things can be, play an important role as well.”*  *“So I tend to give them, I tend to ask them what’s your writing and reading like, and explain why I ask that, and if their reading’s not too bad, or they've got a relative who is a good reader or is interested, I will give them some appropriate literature.”*  *“I try to make it very patient specific if somebody's very clued in and their daughter is a cardiologist, I’m not going to be getting too bogged down on it, not that to be honest, that doesn't happen too much in our demographic. But I do try to you know we all do you know we're very experienced to sort of leaning into what someone's particular needs are”*  *“I tend to give them give them written information, give them as much data as they want, and try and respect their educational attainment and their abilities.”*  *“That's the beauty of primary care. You know they’re health seeking, you know they're not going to come back, or you know that they are. We're good at looking at compliance. We're good at some, you know what what could make that easier.”*  *“There needs to be an appetite for an acceptance that everybody's got a different approach.”*  *“Certainly, if I have patients that I've been, I don't feel they need to have a degree, but I would normally have a good idea of their intellectual capacity. I'm not that interested in their attainment, again, locally, we would have an awful lot of patients who are very capable, who just never got a chance to go to a uni and so I don't use that as a as a yardstick.”*  *“It's just us as a cohort, understanding that the basics need to be done so so so so well but making a provision for your more able individual, so they don't get bored or feel lectured to or feel patronized, but making that type of resource available is fascinating, you know, to see how people learn differently”*  *“Therefore they they they don't want to have a diagnosis of ischaemic heart disease, despite the fact, it's looking at them straight in the face. Sometimes ignorance on on behalf of the the the patients as well, although I’m saying a lot of them know what they they have, there's other patients will, will, will not, or will choose not to to know what what's going on, and so like lack of knowledge as well.”* |
| Limited Processing Capacity | Patients described the experience of not being able to remember or process important information given by clinicians during their heart episode or diagnosis appointment. One clinician also reflected their awareness of this experience and described how they navigate this in their clinical practice to ensure appropriate communication with their patients. This limited capacity to process their diagnosis can have substantial impact on patient understanding and therefore also the efficacy of their treatment. | *“When I first went to the cardiologist she she told me that, she explained that yes, she agreed that it was angina a form of angina I was having, I didn't hear her say that. What I actually heard her say was the complete opposite. And like, what I heard her say was, oh, no, you you you you you're not having angina and now why I heard that, because obviously that's what I wanted to hear. I don't know. But I said oh that's great and she said no you do, no no she said, no, I’m telling you you do have heart disease. That's what I’m telling you. She had to kind of say it to me twice. So for some reason I didn't hear it the first time she said it.”*  *“You can't always be relied upon to remember what's certainly what's been said to you when you're in a crisis like that.”*  *“I think it depends on what state you're in at the time whether you can take in any of that information or not.”*  *“When you go in and you've got you've had a heart attack, whatever they said to you, you don't hear, anyway because all you're concerned about is basically you've had an heart attack, you're either going to live or you're going to die.”*  *“Anybody who's been ill, and I trust we were all in that situation, can probably identify with the fact that when you're ill you don't know you're ill. As far as you’re concerned, you’re fully compos mentis and able but you’re not. You're as mad as a March hare.”*  *“To my warped mind at that point in time, and I fully accept that I I left hospital far too early. I was as mad as a March hare but I didn't think I was.”*  *“It's possible that they did tell me some of this information, but my memory was so bad, both short term and long term when I was in the hospital, and for some time afterwards it might be that they've told me and I’ve forgotten it”* | “Well, I mean certainly my experience with the majority of patients, as I've alluded to, heart disease carries a huge emotional cost to a lot of our patients. My experience as a full time GP in one of the busiest general practices in Northern Ireland is that when you give somebody that diagnosis they are frequently anxious and stressed and I am very aware of the model of cognition that happens whenever somebody gets those diagnoses, so they delete a lot of information, and if I’m honest particularly the women, in my opinion, they distort a lot of the information.”  “What I've learned is I don't tend to challenge them when I say I’m bringing you back for discussion, or whatever, because it's not the truth if they haven't been told if you've documented it well, but it is their truth, and they're not telling lies, they're just deleting and distorting in the context of a complex diagnosis. So that's probably my my my own method, and I think that probably happens throughout the practice.”  “Again, culturally, because it is such a bomb, I tend to acknowledge that even in my own head, even if I don't necessarily say it to them. I have a very low expectation of the amount of information they're going to take on board.” |
| Patient Autonomy | There was a mixed response from patients regarding the extent to which they played an active role in the diagnosis, monitoring and treatment of their condition. Some patients described full autonomy in attending appointments, tracking symptoms and managing medication dosages, while others felt they were abandoned by their doctor and were unable to maintain consistent self-monitoring or communication with health services. Meanwhile, many clinicians expressed concerns with an over-reliance on health services from patients, and the expectation for patients to take responsibility of their own care following the initial diagnosis made by a medical professional. | *“They would observe me for a few hours and discharge me. I think I went to hospital twice, and on the third occasion, I thought, well, what's the point they are just going to discharge me so I didn't bother.”*  *“I was in dialogue with my GP practice. But that sort of ground to a halt during the pandemic for some reason, that they you know they they weren't following up, asking for my readings, so I just stopped doing it”*  *“If I don't know what the norm is for that particular unusual factor then I look it up so that I have a rough guide as to how I’m doing because it makes me feel in control”*  *“Also I can ask questions then, and say, are you worried about this? Should I be doing that? You know, so if your potassium is high, you can adjust your diet accordingly. And you know, if your Warfarin is too high, too low or whatever.”*  *“I purchased myself like one of these blood pressure monitors for at home, and I used to take it almost daily.”*  *“I just took myself to to the GP”*  *“I asked for the recording”*  *“Anything that goes wrong that’s obviously probably my fault for not listening to them.”*  *“My drugs I actually self-administer. So, for example, you know, if if my blood pressure gets too low, I tend to cut down on the drugs.”*  *“I was saying that one thing I've always done is request my hospital notes covering the stay that I was in hospital, and they're very revealing”*  *“Some people sit back and just accept whatever’s thrown at them”*  *“I’m one of these, question everything that's thrown at them, and for me I've got to be able to square circles and I couldn't understand what had caused it”* | *“So some if you said there’s things you can do at home would really engage with that. Others would just be. no, no I just want to see a specialist to be told what the problem is.”*  *“So some if you said there’s things you can do at home would really engage with that. Others would just be. no, no I just want to see a specialist to be told what the problem is.”*  *“Well then the practice can't chase everyone's, you know we're saying we can't chase it, you need to do that yourself, you know we'll tell you the number, but you've got to ring them up and do it and then I mean, yeah, I suppose they're probably some of them not as motivated to do that.”*  *“More useful for a lot of patients as well, if I walk from my house to the bus stop, I stop once. So if you're starting to, you know, if they're noticing they’re now not walking to the bus stop, you know, even stopping once or that you don't need to stop once that’s a good indicator. We also have shown patients the step count on their phones so they can use that.”*  *“People with sort of foot, we're looking at changes in the foot, we might say, take, if you're worried, take a photo and send that to us.”*  *“Also often that they've not thought on their side to bring somebody. So, for example, you know that there are patients who are and this is generalisation, who are young who otherwise completely functional working human beings. Why, they feel they could not have  you know, if they know they're coming to appointment and there's no way they can communicate, perhaps they might have thought about how they might address that as well. ”*  *“I think we need to empower a patient as much as possible and not leave them in a dependent state on the NHS because the NHS cannot, can no longer provide that sort of mollycoddling.”*  *“It would be a patient would make an appointment, either with the presentation that they suspect is heart disease themselves”*  *“there's no active monitoring. The monitoring itself would depend on the patient staying in touch with with me.”*  *“first challenge is getting the patient to actually engage to present.”*  *“Second obstacle is is time, access to primary care itself, and the patient actually managing to make an appointment with me, getting past the the appointment system”*  *“It's now over to them to try to make the changes to their life to to reduce the impact of this going forward”*  *“I think, there’s probably a huge, undiagnosed burden from Covid so we're trying to catch up with that and those patients may be not people who will determinedly approach us for review they may be people who would maybe sit back, and maybe their symptoms, they potentially feel they’re not that serious, but in reality they're probably the potentially the sickest people so so it's it's difficulty in accessing yourselves,”*  *“We've got a big hemochromatosis population, for example, and you're trying to get them back for fasting iron profiles, and you know that's linked to cardiovascular disease, etc. So, I I tend to, you know I diarize those kinds of people, so I keep a kind of close eye on it and and understand that they're not they're not, what I've learned from doing this now for 25 years is it’s very it's it's not very likely that somebody with a cardiac condition or cardiac disease is sitting at home thinking I need to get all this sorted and I just can't be bothered, there's myriad reasons why they don't engage.”* |
| Psychological Impact of Diagnosis | There is clear psychological and emotional distress associated with receiving a heart disease diagnosis that was found amongst the majority of patient stories. Failure to consider the negative psychological impact of being diagnosed with a heart disease can leave patients feeling overwhelmed and alone in the face of their condition. It is crucial for clinicians to take this into account, although only a couple of those interviewed specified ways that they try to acknowledge and support their patients with this specific challenge. | *“When when you're, I won't use the word afraid, but but when you become aware and start thinking”*  *“Or I don't feel great, but can't go to that party, because I just don't feel as though I want to mix.”*  *“It was really talking therapies, really that's uhhh what I needed. But it was important. It was, looking back, quite an important factor in managing the condition up until I had the ablations. Once I’d had the ablations, everything was a whole lot lot better but prior to that, when I was still having symptoms on a more regular basis, it was very troublesome and, and I did need that support.”*  *“During the walk she tries to give you a a pep talk, telling you that there's no more cream cakes, or fish and chips or fry fry ups in the morning. You're feeling bad enough as it is, without getting told what you can't take.”*  *“I found the diagnosis devastating. I was 24 in a hospital, youngest by many years.”*  *“I think that the professionals don't quite realize how much they're frightening the patients before the operation. I think that was my biggest concern.”*  *“Having a heart attack or having a heart episode is a hugely emotional thing…So it's not just definitely not just heart disease but but the emotional impact that does need to be considered.”*  *“I felt that soon as I, when I had the heart attack, my life seemed to stand still. Not just physically, but also mentally.”*  *“I was still young, you know. I got young grandchildren everything, and and I just felt like I'd been you know, given a life term sentence with with mine. So mine's been a gradual I mean my first heart attack was over six years ago, so it's been a gradual process in terms of mental, mental, mentally overcoming it.”*  *“On the subject of overall health and wellbeing things that are important to you that the mental health side of things can be affected”* | *“Well, I mean certainly my experience with the majority of patients, as I've alluded to, heart disease carries a huge emotional cost to a lot of our patients.”*  *“So I do try to lean into the fact that the diagnosis is a bit of a bomb for a lot of people and then I tend to sort of say them right, go away with all that, and then I tend to tell them I'll contact them in four to six weeks.”*  *“I don't, we we we're now part of a multidisciplinary team which to us is great, but to the patients it’s often a push. So if I said to them, for example, right I'm changing this, and I’m changing that, make your appointment with the pharmacist, my experience of that, particularly in our older clientele, is they feel abandoned”*  *“One of the big problems in secondary care is there is a group of colleagues who feel that, the quicker the turnaround time, the better the clinician they are and they fling them in, and they fling them out and the patients are often quite psychologically traumatized by these fast diagnoses”*  *“He was diagnosed with unstable angina, sent to the Cath Lab, got Cath-ed and got four stents and was discharged within 24 hours. He had extreme trauma after that. He made an appointment to come into me, and he said, oh, he called me Dr [CL010], that's what they do, I have 16 questions, and I had to lean into that in a 10 minute consultation”*  *“I think it's about trying to remove the fear from the from the patient trying to de-escalate them, because you, you'll often see panic, particularly in the part of the world that we live in, where there's a very high incidence of ischaemic heart disease and explain to patients that the important thing is making the diagnosis and then making the changes that are needed to reduce the impact of that diagnosis. So the the the management, to my mind, is all about reducing fear to allow the patient to engage in the management of their of their condition.”*  *“I will acknowledge that this has caused upset to to the person in front of me, or to the person on the end of the phone depending on how, how we're dealing with it, and I’ll ask them why they would feel that way or what is concerning them, or are scaring them up about it to acknowledge that, that it may be scary and off putting and just just try to bring them back to the the goals that that I discussed before to try to get them to engage with the fact that they, this is something that they can do that will minimize the impact of this condition on on their life going forward.”* |
| Impact of Socioeconomic Status (SES) | Clinicians describe how patient SES can influence the way they interact with healthcare services and this needs to be factored in during the process of making a diagnosis. One clinician describes in detail how they change their approach to appointments when dealing with patients who have lower literacy rates or limited access to personal health information. Though this was a relatively small sub-theme, there was an important message regarding patient affluence and education influencing clinician expectations and approaches when making a diagnosis. | NA | *“And the the area I work is not the most affluent so often, it's they, they don't even know, even them trying to say to them, you need to go and ask your family about your family history. And do, you know, go and ask whether anyone's got high blood pressure? Have they got diabetes, are your family on statins? That kind of question, just to get a bit more of a a kind of a picture of what's actually happening in their family.”*  *“I think they're probably more tolerant of certain things in some respects.”*  *“I suppose sometimes, from a clinician point of view, it can be easier because I think there may be not as demanding as when I've worked in more affluent areas, and they are more accepting of the wait time. But they may not be as proactive and actually chasing that up and kind of following it through. I mean’s it's not a completely deprived, so you know, most people have a phone, there is no reason they can't ring the local hospital and chase up when their appointments is.”*  *“I do it face to face for lots of reasons, mostly being the literacy issues that a lot of our patients would have. So demographically, I work in a practice of 10 and a half thousand patients. There's 330 practices in Northern Ireland, and we're probably the thirtieth most deprived and obviously with that comes all the concomitant concerns and issues, particularly literacy and health literacy. But never mind health literacy, basic, you know concept of, literacy, is a big issue so I bring them in, and I've got forms and sheets that I go through with them and give them information.”*  *“Big problem for doctors as patients talk in terms of, and obviously again with very poor and poverty ridden demographic, a lot of them roll their own cigarettes, and they say I only take half an ounce, I don't even know what that means. I'm like well, what is that, you know? So I will ask the patient to tell me what that would be roughly in terms of normal cigarettes.”*  *“You're looking at patient background, and their socio-economic background”* |
| Varying Availability of Social Support | Patients shared varying levels of personal support in their lives, with some owing their lives to family members and colleagues who were nearby at the time of or following an episode, while others described the struggles associated with living alone and not having family nearby. This sub-theme highlights how different patient access to support can be and the realities of loneliness in older adults diagnosed with heart disease. | *“I was in work sitting opposite [Name] who is the [Hospital-name] Ambulance Volunteer. And she saw me looking pretty awful and said was I feeling okay? and I said, I wasn't, so she called an ambulance”*  *“But I was a Trust governor at the time and went along to help out at a group and it was, it was run by a nurse who was very interested in different of types of atrial fibrillation and a specialist. I ended up, I missed the first group, and the second one I joined and stayed for years…talks from dieticians and pharmacists and it was the most helpful of any treatments I've ever had and I stayed with that group for years, and unfortunately I think it was the pandemic that sort of stopped everything because they didn't do Zoom or Teams but I would recommend that wholeheartedly, so some bad aspects of it, but an awful lot of good, and I met some great people who are still friends, and we still meet up regularly and chat but I learned a massive amount about the condition from that group”*  *“The local hospital was about a mile and a half from where we were, so they decided to basically throw me in the back of a car and bring me to the hospital.”*  *“listening to all the people who came, and their, like today, their experiences. It kind of made me think, well, mine hasn't been that bad in comparison to what others have gone through.”*  *“In particular my colleague [Name] and who actually saw what was happening and took action, called the ambulance and actually performed the CPR. So yeah. I owe a lot of people a lot.”*  *“I kind of thought for my family that would be dreadful if anything happened or you know, if my husband had to come, he'd be on his own, he’d have no support. I knew I’d be in possibly overnight, so I elected to wait, and as it turned out, it wasn't as long a wait as they anticipated, but I didn't look on it unfavourably. I just thought that was a really tough decision for people to make, you know, especially if they were on their own, maybe.”*  *“My greatest challenge was, I live on my own. And it was just looking after myself because everything was too much effort. Would I say effort? It doesn't sound right saying too much effort. But just washing a cup for a cup of tea would would send my heart into distress, you know, the strain of it. And and there was no support”*  *“Three years later, I collapsed at home, unconscious and luckily my son was there and put me in the W position and take me to hospital,”*  *“I think the only persons that that helped me was family. And that's that's all I can say is family got me through it.”*  *“As an expat you don't really have the right network of family members over there when you are living abroad, anyway. So that's difficult.”*  *“My wife dialled 911 uh 111 and they freaked out and said that the paramedics had saved my life.”*  *“What saved my life was my brother walked into into the house the second that I crashed. He began CPR.”*  *“What I have done is, I detest Facebook with a vengeance, but I have found a very good support site on Facebook, called SCA, which is sudden cardiac arrest, where, in order to qualify for the group, you have to have had one, and I've learned so much from that dealing with memory issues, cold issues. You you name it. It it appears that there's many, many, many different reasons why people all have the same thing.”* | NA |
| CL Clinician. P Patient. Redacted text shown in []. | | | |

| **Theme 3: Patient-Clinician Interactions** | | | |
| --- | --- | --- | --- |
| **Subthemes** | **Description** | **References** | |
|  |  | **Participant Experience** | **Clinical Experience** |
| Communication | Patient experience of communication with clinician varied considerably between participants, with some feeling satisfied with the degree to which information was explained to them, while others describe feelings of confusion and lack of understanding of what was happening to them. Majority of clinicians acknowledged the importance of communication with patients, however, they also expressed concerns with time restricted appointments preventing them from being able to adequately communicate with their patients. | *“But it's a concern. It's the uncertainty of what really happened because it doesn't quite get explained to you”*  *“I remember them discussing Warfarin, and why it was important in the chart that you were assessed, if you were a woman, if you had a certain age, you know risk factors. So I remember them going through it”*  *“I mean, I've had people ringing me up talking to me on the phone, and then they write a letter that the Royal Mail, deliver it in the post, and what is all that about because they have never met me, they know nothing about me!”*  *“We talked about diets and technology and fitness and things like that and touch wood since then everything's been okay.”*  *“The heart surgeons back then were very matter of fact, and I was just told this is what was happening, and you'll be fine there was no talking about it like there is now”*  *“Similarly, pretty young I was like, what on earth is going on?...There was no discussion about anything else, just either go on medication or have ablation. So I waited for ablation till 2002.”*  *“So my obviously mine was only 12 years ago, but the team that looked after me were absolutely fantastic, and they talked about everything, and you know we came to, you know, disagreements and agreements.”*  *“Obviously in social work, I’m used to dealing with medical professionals a lot, I really valued all the information that I was being given. I really valued the fact that when I went down to have my first lot of stenting done.”*  *“I valued the fact that they came in each of them came and told me what they were.”*  *“Just not much communication. I've got a very good consultant. But yeah, it's it's the lack of information”*  *“I think, better monitoring of people with no risk factors and also better information given to them, like when I was diagnosed with diabetes, as I said, it was two years, two, three years before, three years before I got my stenting done, there was no discussion, I I mean, I now now, I kind of knew, anyway, but I know that most people probably wouldn't, I know now, there's a huge connection between heart disease and diabetes, but it was never discussed”*  *“The hardest part is, deliver it in a way to the individual not to stress them out but again it's it's probably putting too much on the GP in the end.”*  *“The hardest part is, deliver it in a way to the individual not to stress them out but again it's it's probably putting too much on the GP in the end.”*  *“To be fair the staff there are absolutely fantastic. They are really good at explaining things”*  *“But certainly, I have no contact with my doctor,”*  *“And just having that like really open and frank conversation was really useful for me personally. So that was the most important thing for me”*  *“Basically, I had no information whatsoever”*  *“At the time of of diagnosis, I think, there is an essential lack of information from the doctors.”*  *“I didn't really get any reassurance from the doctor about how bad my condition was, or you know at what stage it was in in the process”*  *“I think that that that was the worst bit for me when I was first diagnosed. I didn't just didn't know what the future was going to hold or how long I was going to survive.”*  *“For me they showed me a picture of a before and after of my artery right after my surgery so I could see the difference…It was useful as I could see that my artery was more open”*  *“There's one or two hidden gems that nobody ever ever told me about, and I wouldn't know about, unless I’d requested the notes…I learned a lot more from having read through those notes than I ever did from being told anything, either pre, in, or post care.”*  *“It was very, very difficult to get any communication with [Hospital- name] because they were just too busy.”*  *“It has to be better information, as I said earlier, general information out there for people, you know. I've seen signs on for GPs about about heart attacks is actually, the ones I said about pain in the back back, pain in the arm. But you know there needs to be better better communication about what symptoms might be”* | *“I think you start with the patient you would normally the patient would like to know*  *you know some details about the condition they have. You know, the heart condition or the vascular condition they have and what exactly they have. What are the you know, normally, I explain you know the mechanism, the cause, you know the underlying cause of the problem, how severe it is. And then you know what’s the prognosis, what is the, is it something that is long term or something that happens only for a short time. Is this something curable? Is it something that can be treated easily? Or is it something that cannot be treated, you know, cannot be cured and what are the treatments available and how how are they gonna affect their lives? so all these things are important, so so so the patient, I think most patients would expect you know some explanation on all these factors.”*  *“I think communication is extremely important in medicine. It doesn't matter which field of medicine. In fact since medical school a lot of emphasis is put on communication skills because at the end of the day the the in medicine is about, it's an interaction between the patient and the doctor. You know, it's not just making the right, first of all to diagnose something correctly, the communication of getting information from the patient is important and then how are you going to deliver the news, the treatment plan to the patients as equally important as well. I think communication is is a key to the whole management or the care to the patient.”*  *“It could be a cultural barrier. It could be a personal reason as well. Some clinicians may have personal reasons you know, difficult to express specific things. Yeah. So so this are the from the clinician’s side.”*  *“I think you know going back to basics like training is important. It's the you know. This is why medical school communication skills, you know it it starts from day one even up to you know someone becoming specialist, even at my stage when I’m fully trained you know I'm still learning to communicate better.”*  *“Facilities will help, you know, tools to help with the communications like drawings, pictures, things that would be able to help, which can inform the patient better about the condition and then having enough time.”*  *“I think time is a challenge in the NHS. You know, in this case I'm only given 10/15 minutes to see one patient, which I don't think is enough, you know, giving, you know we need to have a you know, some conditions, especially in the complex ones will require you know a long, you know, half an hour, an hour time to communicate properly with the patient.”*  *“The lang, you know culture again, cultural language barrier, all these things. Some of it you cannot address like if I can't speak your language, I can‘t but having an interpreter, it helps, but it doesn't it doesn't address everything. But again it’s one of the skills that can be tried to learned through training and experience.”*  *“So communication issues, and you know we do have interp so say language. We may have interpreters but very often that is, you know, there are still limitations with that. And so, yeah. So communication in an already limited or timed setting.”*  *“Well generally it's um it's I think it comes with the experience. So now, having done it for a long time, you kind of can gauge better of how people are taking in things, I think and recognising that everybody is very different. So how I might have a conversation with one person or one family might be quite different, based on how you're reading their feedback to it, and I think I found that easier as time goes with experience, I think ”*  *“patients coming who clearly don't speak the language, or for whatever reason cannot communicate and we've not been made aware of that. So either the referrer hasn't told us or we've not, either I don't know, found a way of getting help, or indeed, even trying to book a longer slot or also often that they've not thought on their side to bring somebody. So, for example, you know that there are patients who are and this is generalisation, who are young who otherwise completely functional working human beings. Why, they feel they could not have  you know, if they know they're coming to appointment and there's no way they can communicate, perhaps they might have thought about how they might address that as well.”*  *“Then the key thing is the history. Speaking to the patient. Letting the patient often for firstly, for an open question let the patient speak for themselves, describe any symptoms, try not to interrupt them during this period and from there you sort of go from the open questions down to some more sort of closed questions”*  *“So I think the you know the the patient's opinion I think, is first of all, it's most important. What they feel is going on. What they're worried about. These things are, you know, so it's an important to involve the patient at any point. ”*  *“So obviously you want to keep them and their families as well informed to what's going on, reassure them, tell them you know what you’ve found. You know what you think’s going on and being honest with them as well. If they’re unwell then letting them know”*  *“In fact, if we do get come to a diagnosis or a a temporary diagnosis then let them know what their diagnosis is, and also what the plan is from there on and let them know the the risks and with this the benefits of treatment, and then the risks of treatment as well, keep keep them well informed throughout the the the process.”*  *“You know any suggestions about what with what resources we've got here we could do to make help and aid in the diagnosis as well. These I think that’d be that'd be that'd be useful. ”*  *“Be it as I say, a rhythm issue, I tend to describe it in my head, are they a plumbing issue or an electrician’s issue, and that's probably the best language I can use with patients, because they find it very difficult to understand the difference frequently between atrial fibrillation, for example, or heart failure, or ischemic heart disease”*  *“But I do try to you know we all do you know we're very experienced to sort of leaning in to what someone's particular needs are”*  *“I tend to try and bring them back, tend to understand that they're not going to remember it all. Probably one of the major challenges as a clinician is whenever someone comes in and says, no one told me that, you never told me that, and I have had to do some work on that myself, because you're sitting with the information in front of you. And what I've learned is I don't tend to challenge them when I say I’m bringing you back for discussion, or whatever, because it's not the truth if they haven't been told if you've documented it well, but it is their truth, and they're not telling lies, they're just deleting and distorting in the context of a complex diagnosis. So that's probably my my my own method, and I think that probably happens throughout the practice.”*  *“This is our poor communication. You know your swab was positive. People often means that they think that that means that there is nothing growing on it, but in actual fact it’s positive for something. One of the worst scenarios is an oncology patient is told your scan is positive, and they think that's positive news. They go into deletion and distortion. So it's just us as a cohort, understanding that the basics need to be done”*  *“I think communication with the patient is is imperative”* |
| Continuity of Care | Patients expressed feelings of frustration in regard to the lack of continuity in their care, especially older participants who have memories of an established family doctor who they were familiar with and felt known by. While clinicians also acknowledged the patient frustration, they saw the current system as beneficial overall and believed patients should be supported in adopting this approach to providing healthcare services. | *“Even though I've been diagnosed, I still kept getting ad hoc checks through NHS-provided tests”*  *“But since then you know the care from the GP is horrendous, that you know it's me chasing all the time where you're supposed to be on six month reviews for certain things and three months reviews”*  *“I think I think in this world, you're lucky or you're unlucky so you know I’m just unlucky with with the surgery I’m with and I was very lucky with the consultant I had.”*  *“It's lack of access to one GP, so lack of access to the same GP. My surgery has just joined, not just, about two years ago, joined with two others. And now if you, if phone to speak to a doctor, you invariably get somebody you've never spoken to before and I think that's you know the continuity of care is reliant upon very good recording by GPs, which I think doesn't always happen.”*  *“On return back to the UK, when I went back to my GP because I'm in a new city. Basically, the the locum doctor was excellent, but unfortunately, she left after 6 months, and I've had no follow up treatment since.”*  *“That's all they said, I'll ring an ambulance and the ambulance will come out to you. Four hours I was in the ambulance and 26 hours I was in the corridor on the trolley. And since then nobody's been interested in me.”*  *“As I said doctors are good, I get the impression that they are brilliant when you are in critical condition and after that, I don't know they seem to forget about you or I don't know. I don't really know what the problem is, I I just can't understand why then they don't care any longer.”* | *“I think probably just with how general practices at the moment actually following up the patient, and then seeing the same person again, is quite difficult really.”*  *“so they, they may well never see the same doctor again, because it's quite a big practice, and there are 10 doctors and three or four advanced nurse practitioners, so they may often in all honesty, not even see the same doctor again.”*  *“I think it's it's probably quite frustrating for them isn't it because they feel like if they saw the same person, they wouldn't necessarily have to keep going over the same things again that actually if you know, actually, if it was just one person who dealt with it.”*  *“I guess that isn't the model in general practice, unfortunately, that they may well see doctors in the beginning, and then they're probably seeing health care assistant who might take their bloods and blood pressure, and then they might see a nurse who goes through those results with them, and I mean, even if they they are being followed up in clinic, they're often not followed up by the same consultant that they saw initially.”*  *“now not only do they not have necessarily a a a doctor that knows them well, the doctors, the GPs, don't have time to do this”*  *“Well there, there are no interactions. So so once the patient is referred until they're seen in clinic, and there's probably no interaction with the patients, once they're seen in clinic, then we will arrange some tests, perhaps, and then we would write the patient with the results of those tests, and the need for any further tests or review. What we're trying to do is have very much a one stop shop. So patients are seen, diagnosed, and treated or discharged.”*  *“I don't, we we we're now part of a multidisciplinary team which to us is great, but to the patients it’s often a push. So if I said to them, for example, right I'm changing this, and I’m changing that, make your appointment with the pharmacist, my experience of that, particularly in our older clientele, is they feel abandoned.”* |
| Cultural Influence | Although this was a small subtheme, a few clinicians did suggest that there are cultural barriers between patients and clinicians that may affect how they interact and ultimately the quality of care clinicians are able to provide or that patients feel they are receiving. | NA | *“It could be a cultural barrier. It could be a personal reason as well. Some clinicians may have personal reasons you know, difficult to express specific things. Yeah. So so this are the from the clinician’s side.”*  *“The lang, you know culture again, cultural language barrier, all these things. Some of it you cannot address like if I can't speak your language, I can‘t but having an interpreter, it helps, but it doesn't it doesn't address everything. But again it’s one of the skills that can be tried to learned through training and experience.”*  *“Maybe cultural, personal, whatever it is so sometimes, on some occasions people might um uh not give you all the information. They might choose not to. They might not, you know, and and it might not affect the diagnosis as such, but it might affect how you might go about discussing or agreeing to a treatment plan. So if somebody thinks that if they tell me they have certain symptoms, I'm more likely to go down a certain route or not then you know, that obviously affects it but it may not affect the actual diagnosis because let’s say, we realise that but it obviously affects the conversation.”*  *“In Northern Ireland culturally, the heart, hearts are a big issue, heart disease are a big issue and people often find it difficult to differentiate.”*  *“We have to be respectful of the fact that as I say, culturally in [City] heart attacks, I mean Benecol is like, you know, the the Holy Grail. You can, you know, go into any supermarket, and it's full of these, you know cholesterol lowering agents.”* |
| Expectation Management | There seems to be a disconnect between patient and clinician experiences when it comes to expectation management, as patients described frustration with the lack of reassurance or explanation from their healthcare provider, while clinicians from our sample described how they support their patients by keeping them informed and explaining the chosen treatment plan. While this may be the result of a biased sample, it could also speak to an existing disconnect between patients and clinicians during the process of diagnosing a heart disease. | *“The heart surgeons back then were very matter of fact, and I was just told this is what was happening, and you'll be fine there was no talking about it like there is now and just I was just expected to accept it really which was tough.”*  *“It was just really useful to sit down with someone and just chat about it, was was really useful for me because I had a lot of like questions, and I was like, don’ t really know what I'm doing, like. What what should I be doing? What should I be like keeping an eye out on?”*  *“I didn't really get any reassurance from the doctor about how bad my condition was, or you know at what stage it was in in the process, and in fact that's in very, very extreme cases that you know that death is going to occur within two years. But at the time when you are sort of under duress anyway, after having had this diagnosis you tend to sort of go to any point of knowledge or information. And I think that can be quite dangerous, and that you know, because your your situation and your diagnosis is very personal to you and you, you know whereas you go on the Internet. It's looking, it tends to sensationalize a bit, and you know it'll be the worst possible scenario.”*  *“I think that that that was the worst bit for me when I was first diagnosed. I didn't just didn't know what the future was going to hold or how long I was going to survive.”*  *“I think any like additional steps that the clinician, or or the person themselves needs need needs to do as well would be probably useful instead of just being given like here is, here's the situation, and it's like no real like next steps. I think any sort of like helpful next steps is always like quite useful, even if it's like you'll see this person in like a month or two, or something like that”* | *“Being able to give them a timeframe…I think if we could have that prompter for patients, they would actually kind of feel more reassured by that”*  *“I think they're quite frustrated, because often they'll book in to see a doctor and they'll say, oh, I saw someone two weeks ago, and that letter, because of all the backlogs hasn't been typed up and arrived or you've had one bit of paper with the medication they want prescribing, and then they said, oh, you know I'm supposed to come in in a couple of weeks to see whether it needs changing, but we haven't yet had that information through. So I think to be honest, a lot of them are quite frustrated, you know, and you can understand why, because from their point of view, they think, why, why, in this day and age isn't it an email just sent, and that's available? I suppose they, you know, they kind of they don't understand necessarily the process of what's happening”*  *“I usually will say to them that it is unfortunate that I have to be frank, honest, and able to disclose everything”*  *“I think you start with the patient you would normally the patient would like to know*  *you know some details about the condition they have… what exactly they have. What are the you know, normally, I explain you know the mechanism, the cause, you know the underlying cause of the problem, how severe it is. And then you know what’s the prognosis, what is the, is it something that is long term or something that happens only for a short time. Is this something curable? Is it something that can be treated easily? Or is it something that cannot be treated, you know, cannot be cured and what are the treatments available and how how are they gonna affect their lives? so all these things are important, so so so the patient, I think most patients would expect you know some explanation on all these factors.”*  *“So as a clinician when I first meet the patient I keep my mind very open and try to gauge how you know their background, their expectation”*  *“It's very important the patients are happy when they leave the clinic I mean vast majority of people are, because there's a plan, even if it's not good news, there's there's a plan for what we're going to do to make them feel better. We're very lucky in cardiology, because many patients come with symptoms, and we're able to offer them a range of treatments to make them feel better or to reassure them there's not a problem”*  *“In fact, if we do get come to a diagnosis or a a temporary diagnosis then let them know what their diagnosis is, and also what the plan is from there on and let them know the the risks and with this the benefits of treatment, and then the risks of treatment as well, keep keep them well informed throughout the the the process.”*  *“I try to do as much of it as I can, attach as much information as I can, have honest conversations about the fact that they may just require an investigation, like an Echo for example. And I will always write that in the letter, this patient's happy to proceed to investigation…I tend to sort of say them right, go away with all that, and then I tend to tell them I'll contact them in four to six weeks”*  *“So I will often be very prescriptive at the start, particularly with the ones that are a wee bit more able “I'm going to be seeing you reasonably regularly and then I'm going to be extending that out and then you're gonna be finding your own feet” and that is, I need to make them aware that the proactive follow up is not a long-standing issue, I can't be in a long standing provision because I have more coming up the back that needs to be sorted, and I'm really keen from the outset to let them know when I let you go or I pass you on to an MDT that is an absolute sign of progress, and it's a sign that you're going in the correct direction.”*  *“He said, oh, he called me Dr [CL010], that's what they do, I have 16 questions, and I had to lean into that in a 10 minute consultation. To be fair, a lot of them are very straightforward, and then I was able to explain to him our process, and how we follow that up, and the drugs are on the repeat and we've got you down for a six week review, and we've got you down for three months for your bloods, he was very reassured”*  *“If I’m giving anybody something to do and they manage it themselves, I'm very clear. Do this during the week, don't do it on a Tuesday, because I’m not there. Be, be sensible, and do this when you can get some support and feedback from people that know you the best.”* |
| Patient Experience with Healthcare Systems | This subtheme highlights the wide range in patient experiences with healthcare services. The data included here suggests that quality of patient care are not constant across individuals. Clinicians have a varying degree of awareness of these differences although one interview directly expressed that patient experiences will depend on the clinician they are dealing with. The data implies a degree of luck in having a positive clinician experience | *“I was diagnosed as having a murmur at the age of four, but that gradually became, the the story changed over the years, a hole in the heart, aortic stenosis, you name it. I was diagnosed with it. It's a, I mean I'm 66. So we're talking about over 60 years ago that there weren't all the ultrasounds and that sort of thing, so that they were guessing much of the time”*  *“When I was discharged from the hospital, they gave me a bag with some medications in it and a letter from my family GP. The following day, a family member took the letter down to the surgery and the following day after that, I got a phone call asking me to attend at the surgery later that afternoon.”*  *“I mean the people that stand out for me ironically, my GP, even though he misdiagnosed the heart attack as as a bad indigestion. Obviously the medical staff at two hospitals [Hospital-name] and [Hospital-name], who were quite brilliant,”*  *“I hate to say this because I I shouldn't have to say this. But nobody that I encountered in the NHS helped me. In fact, some went out of their way, and it might be because they didn't like me. I I can be an abrasive character but some of them went out of their way to make life difficult for me. When I was in hospital, the nurse would give me furosemide to make me urinate and she would disappear and I was left there with a full bladder, not being able to leave the bed and not being able to empty my bladder. The doctor said if the if he's in pain give him pain medication, and it was written on the board, and yet the nurses refused to give me pain medication! Why would they do that? It was I mean, I’m sorry, but it was at best it was people just going through the motions, just getting through the shift and going home.”*  *“Throughout all my different types of experiences with the NHS I have nothing but praise for them”*  *“I had a a nurse that would ring me. She rang three times. And it was only after a while I realized that her sole objective was to try to get me to take more drugs. That was all she wanted to do. I said, I'm taking loads of drugs. How many more do you want me to take?”*  *“I went into GP, and luckily I was seen by very astute newly qualified GP. Because no one had ever heard my arrhythmia, or not my arrhythmia I have that as well, but my murmur”*  *“Once they start of course they don't stop, you keep on getting different medicines and medications, until they find the right one that works or they hope is going to work. You know they just took me off one drug now and partly because it makes my ankles swell but they’ve increased another one and we’re waiting to see what happens with that and I've been, I had the heart, the valve replacement in 2016, and they’re still fiddling with medicines.”*  *“I think I think in this world, you're lucky or you're unlucky so you know I’m just unlucky with with the surgery I’m with and I was very lucky with the consultant I had.”*  *“in '98 you didn't get any of that. You were just taken down for surgery. So we've come so far the other way. I didn't know what the hell was going on. I wasn't even really clear what surgery I was having. You were just taking away and that was it. So things have got to find a middle ground really.”*  *“With the with the second episode, the heart attack, the biggest issue around that was that I had no empathy whatsoever from the call handler and I I that I have actually done a complaint, and that's you know that's all been sorted out now”*  *“Could I say that the staff are absolutely fantastic? All NHS staff are absolutely fantastic. I didn't meet anyone that was horrible, nasty, or anything else”*  *“I was admitted to hospital a couple of, few times actually. Actually, the care when I was admitted was great. They looked after me really well,”*  *“Whilst I was in hospital, the the nursing care was fantastic. I mean, you know, can't can’t fault it.”* | *“Because our literally, you know, absolutely obscene waiting times, I have to lean into that that and try and make the patient’s experience as safe as possible.”*  *“Depending on the consultant they get, they will feel very informed and very supported or they will equally feel very judged and dismissed”* |
| Patient Information Comprehension | The degree to which patients understand information related to their diagnosis varied, with some feeling confident and others feeling confusion. Clinicians acknowledged this challenge and shared techniques they use to support their patients in improving their understanding. | *“There was only one figure that was important to me, and that was a heart rate of 198 beats a minute, at which point the heart is not really pumping blood, its more sort of quivering like a jelly.”*  *“The only number he's mentioned over all that time was with regard to left ventricle ejection fraction and it's not really a number, it's less than 15%, whatever that means. I don't think it's good.”*  *“it's based on your knowledge. So I take my blood pressure in the same environment the same same time every week. So, and I understand what it's going to be.”*  *“but I would say, especially in that initial diagnosis they kind of just gave me a leaflet and they were like here, here’s the information. So, I was actually like initially a bit confused”*  *“But I would say, especially in that initial diagnosis stage I was a bit confused and I was like, trying to just Google information. And obviously, you know, when you start googling information about what's wrong with your heart, usually that tends to be a bit exaggerated, so I probably didn't get the most balanced explanation to begin with and I definitely did ask and yeah, I probably didn't get the time that I felt like that I needed”*  *“I've been given all the warnings and all the rest of it”*  *“I didn't understand the difference between a cardiac arrest and a heart attack. And it's amazing when you talk to people. I think an awful lot of people would be in the same boat, they think they're 2 of the same things and they’re not obviously. They're very, very different.”*  *“I couldn't understand what had caused it. I couldn't understand what they done to really fix it, and I how I would be going be going forward.”* | *“Facilities will help, you know, tools to help with the communications like drawings, pictures, things that would be able to help, which can inform the patient better about the condition and then having enough time.”*  *“I think often the relationship with the healthcare professionals makes a difference, so if they feel that you've understood their problem, and addressing what matters to them, then they're more likely to engage or they've understood the problem.”*  *“Be it as I say, a rhythm issue, I tend to describe it in my head, are they a plumbing issue or an electrician’s issue, and that's probably the best language I can use with patients, because they find it very difficult to understand the difference frequently between atrial fibrillation, for example, or heart failure, or ischemic heart disease”*  *“In Northern Ireland culturally, the heart, hearts are a big issue, heart disease are a big issue and people often find it difficult to differentiate.”*  *“So I tend to give them, I tend to ask them what’s your writing and reading like, and explain why I ask that, and if their reading’s not too bad, or they've got a relative who is a good reader or is interested, I will give them some appropriate literature.”*  *“despite 20 years of really interrogating myself found it really - people get mixed up in percentages. What is a percentage? You know we we all talk in this lingo…this is our poor communication. You know your swab was positive. People often means that they think that that means that there is nothing growing on it, but in actual fact it’s positive for something. One of the worst scenarios is an oncology patient is told your scan is positive, and they think that's positive news. They go into deletion and distortion. So it's just us as a cohort, understanding that the basics need to be done so so so so well but making a provision for your more able individual, so they don't get bored or feel lectured to or feel patronized, but making that type of resource available is fascinating, you know, to see how people learn differently”*  *“They don't know the difference between poorly controlled hypertension, white coat hypertension, malignant hypertension - and these are clinician clinical nurses. If I can't get that through to them despite between 8 and 10 didactic learning courses that we’ve done with them, patients aren’t going to get that. I I I you know, so they in my mind most patients don't get it…So it's easy for people who are in the know to know what the real risks are, but never underestimate how little anyone outside the medical profession gets that…So you know the the health literacy around cardiovascular risks is poor in general, so we need to keep it as simple as possible”*  *“It's raising awareness, and in the population generally about ischaemic heart disease and and reducing the fear that it's sort of reassuring people that things can be done about it, again great strides being made with that regards locally, and the hospital does try to to engage. But that's the that's the first obstacle.”*  *“I think, if the patient doesn't buy into this, you cannot expect them to to, particularly nowadays, where where somebody may even have heart attack and be out of the hospital the next day, having their stent in place. They they don't realize that they do have a serious condition, and it's actually trying to help them buy into that and and realize that a low, the impact so far has been very little, but in previous times this could have been much more significant”*  *“It’s about raising the profile of ischaemic heart disease in in the public consciousness. I I it's similar to the FAST campaign I suppose with with the strokes and CVAs”* |
| Patient Trust in Clinician | The extent to which patients trusted their clinician to approach their care in the appropriate way varied across focus group discussions. Some patients felt let down by the doctors who handled their care and did not trust that their best interests were at the core of their service, while others praised healthcare staff for being knowledgeable and experienced in their field. Clinicians did express difficulties gaining patient trust at times and explained how this could affect the interactions and ultimately the outcome for the patient. | *“One of them a misdiagnosis. Not that I blame the doctor, because you know mistakes get made”*  *“They've actually taken, stopped two of them. So why couldn't they have stopped the 2 of them before?”*  *“He certainly did a lot to save my family from me dying when my sons were under ten. So he is a little bit of a personal hero to me”*  *“Let’s not get caught up on funding, I'll simply say you can't find the problem unless you're looking for it, and I’m not convinced that they've been looking in my case,”*  *“You go on blood tests when you look at the blood test leaflet, and it's not actually checking the stuff that they’re supposed to be checking for, then you have to go back and question it. And you think to yourself you know who's the professional here?”*  *“I don't want to, yeah, I didn't want to have surgery that was so incredibly dangerous. So I chose, or I asked to have it with no sedation, which was a very silly idea. But at least then I had a kind of we had an agreement that he could, he would stop if it was going to be too dangerous an area to work on.”*  *“You're in the ends of the experts. Over the 40 years I've been dealing with them you get to realize that they know 100 percent more than you do so it's common sense to accept what they are saying…So whatever they do you accept, because you know, the alternative is the end…I've met so many of them over the years and I have nothing but confidence in what they're trying to do”*  *“The first thing he said to me is, we're going to keep you alive, which I thought was that's a great thing to say. No, I was really pleased because I definitely knew I was in a bad way, because he said, we're taking control of it now.”*  *“But I have no faith in them doing this, I am afraid.”*  *“I'm still on the same level of warfarin which sort of beggars disbelief somewhere along the line because in theory well, I’m not quite sure how it's supposed to work and I, when I asked the surgery, they weren't quite sure either. So, it seems to be Sod’s law. If it works, keep going and if anything changes, the whole thing will change. It seems a bit ridiculous to me.”* | *“I don't hide things from them but the reason for that is not to frighten them, even though they may well get frightened, but my idea is, you don't win the patient trust until they trust you. So, they cannot trust you until they trust you. And they will trust you when they realise you are probably trustworthy and they will trust you, in fact, more when you tell them what they do not want to hear.”*  *“The patient needs to be you know, to be willing to work with the clinicians and if the patient refuses to work with the clinician then no matter how hard the clinicians try you know it’s not going to work. Again, it's the same factors like culture and and the the education of that. So all these things are barriers but both of them should be able to address if the clinician has experience and has the flexibility to deal with those things.”*  *“Maybe cultural, personal, whatever it is so sometimes, on some occasions people might um uh not give you all the information. They might choose not to. They might not, you know, and and it might not affect the diagnosis as such, but it might affect how you might go about discussing or agreeing to a treatment plan. So if somebody thinks that if they tell me they have certain symptoms, I'm more likely to go down a certain route or not then you know, that obviously affects it but it may not affect the actual diagnosis because let's say, we realise that but it obviously affects the conversation.”*  *“So, for example, again, belief that, you know there might be a reason why they are not opening up and telling us everything. So by the time they come here they understand that we're trying to help. Not so, there's no point, you know, we're not judging, for example or and now not only do they not have necessarily a a a doctor that knows them well, the doctors, the GPs, don't have time to do this”*  *“Making a definitive diagnosis, getting a definitive treatment and responding well to that treatment are probably a key. So patients will, if patients have a good outcome in terms of feeling better, or or a good diagnosis, or something that we fix, then they'll forgive us for making them wait a bit.”*  *“It's very important the patients are happy when they leave the clinic I mean vast majority of people are, because there's a plan, even if it's not good news, there's there's a plan for what we're going to do to make them feel better. We're very lucky in cardiology, because many patients come with symptoms, and we're able to offer them a range of treatments to make them feel better or to reassure them there's not a problem”*  *“I will lose credibility with my patients if I tell them drinking more than two units per day is harmful drinking, so I have to, I have to nuance the conversation. I try and advise them of the impact of alcohol on their heart, and particularly the binge drinking that is part of our culture… so I’ll be doing work around that”* |
| Role of Support and Rapport | Patients shared differing experiences when it came to the type of support they received, however, there was often a strong emotional aspect to both the positive and negative experiences. It was clear that the role of support during the diagnosis process should not be ignored, as it has a substantial impact on patient experience. Clinicians also described the importance of patient-clinician rapport beyond simply communicating the medical information, highlighting how sensitivity and empathy are crucial when dealing with heart disease diagnosis. | *“Prior to this, I hadn't been to the doctor’s for about 10/12 years and I I went, and spoke for forever”*  *“He certainly did a lot to save my family from me dying when my sons were under ten. So he is a little bit of a personal hero to me”*  *“again the cardiologist, just like HD002 has just said, he was really down to earth and he, he kind of said, this is the thing to do, is to have the ablation. You'll get your life back, and you know he gave some really sort of ordinary, good advice.”*  *“the cardiac nurses they they are angels.”*  *“I mean I I'm. I'm amazed that these people are getting support. I was, I was abandoned… there was no support”*  *“The heart surgeons back then were very matter of fact, and I was just told this is what was happening, and you'll be fine there was no talking about it like there is now and just I was just expected to accept it really which was tough.”*  *“I had a nurse put at my head he scratched my nose and things for me, and I tell you that was like I mean, it sounds like such a trivial thing, but it was like a real godsend when you've got your your two arms like this, you know, and you can't move either of them. So I really valued that,”*  *“I had the same, I just happened to have the same cardiologist. She happened to be on, the one that did my work 7 years before, and she pretended she remembered me I mean obviously she didn't remember me, but she'd seen in the notes my notes she had done the work before, and the fact that she came across as somebody who remembered me was fantastically reassuring, when you're in that when you're in that kind of position,”*  *“And now if you, if phone to speak to a doctor, you invariably get somebody you've never spoken to before”*  *“I had absolutely no problem, the ambulance turned up, took to me to [Hospital-name]. Luckily there had been a meeting in casualty, Professor [Name] and Professor [Name], they were all in casualty at the time I think, just having a meeting. They were all going to go home, I think, but they decided to come and see me instead”*  *“Specialist that really knows you knows you and you've met them, and it's not an anonymous. I don't know it's probably asking a bit much.”*  *“I would say like for the most part, the NHS were like pretty supportive,”* | *“ I think my style may not necessarily be acceptable to many but I tell the patient everything, especially if everything is bad. So I don't hold back. I usually start by taking matters seriously, despite my tendency to at times from time to time, you know, cracking a little joke that may or may not be palatable. I also remind the patient indirectly that I am a fallible human being like them and that any statement made by me about their habits or themselves, or their status cannot and should not be colouring my judgment. And, I usually will say to them that it is unfortunate that I have to be frank, honest, and able to disclose everything”*  *“And in those circumstances, if the patient was too upset by the outcome, you may find that I will say to the patient please forgive me for being honest with you, my aim is to help you reach the right diagnosis and right management plan irrespective of whether my intervention is or is not going to prolong your life and I think being truthful, open, and honest with the patient, allows you to then gain their trust in a manner that would allow you to more easily persuade them to do things that you judge to be important for themselves.”*  *“I think often the relationship with the healthcare professionals makes a difference, so if they feel that you've understood their problem, and addressing what matters to them, then they're more likely to engage or they've understood the problem...For example, be it either understood the implications of the diagnosis, or maybe even had a reassuring conversation…if they feel that you've listened to them and explained to them the risk benefit of, for example, an operation they’re more likely to say, okay, an operation is not the easy answer necessarily. So, I guess, understanding the implications of the diagnosis to them.”*  *“you have a very short time to understand the patient, their concerns, their daily lives, and their expectations and hopes and dream isn't it, is all within that so we have quite a short time to do that.”*  *“I think some things are quite simple and used to be done and could just be having more time and building up a relationship. And I think a lot of that is lost in modern medicine, or or or the normal way of working. So you know I, you know not only in the past you might have a family doctor who knows the patient quite well. So they have explored some of these things with the patients already”*  *“During COVID we found this because we thought, can we make use of some of these things? But what a lot of the patients said was missing actually was just not, the the sort of more direct contact.”*  *“making sure that every time before I leave the room, just ask them if they've got any questions, or is anything else I can help them with.”*  *“I try to do as much of it as I can, attach as much information as I can, have honest conversations about the fact that they may just require an investigation, like an Echo for example. And I will always write that in the letter, this patient's happy to proceed to investigation”*  *“I don't send them to Google, I don't send them to Facebook group chats, because then I think that contaminates the quality of what they get.”*  *“And again they're at a time of vulnerability, and you have to nuance those consults because if you hit them up in the face on the first day that you didn't take their cholesterol tablets when I told you, you didn't, and you didn't stop smoking, and you didn't stop drinking, then you just lose them at hello. So you have to nuance it.”*  *“He was diagnosed with unstable angina, sent to the Cath Lab, got Cath-ed and got four stents and was discharged within 24 hours. He had extreme trauma after that. He made an appointment to come into me, and he said, oh, he called me Dr [CL010], that's what they do, I have 16 questions, and I had to lean into that in a 10 minute consultation. To be fair, a lot of them are very straightforward, and then I was able to explain to him our process, and how we follow that up, and the drugs are on the repeat and we've got you down for a six week review, and we've got you down for three months for your bloods, he was very reassured”*  *“I tend to be quite a a chatty person when it comes to interacting with patients. I like to keep things on on a personal level, to try and engage them better in their own management of their of their health. So I tried, as I said before, to reduce the fear, to try and get them to buy into looking after themselves that this is their own health that they're looking after. And by doing this properly, we we will reduce minimise the impact of any diagnosis that they're receiving.”*  *“I will acknowledge that this has caused upset to to the person in front of me, or to the person on the end of the phone depending on how, how we're dealing with it, and I’ll ask them why they would feel that way or what is concerning them, or are scaring them up about it to acknowledge that, that it may be scary and off putting and just just try to bring them back to the the goals that that I discussed before to try to get them to engage with the fact that they, this is something that they can do that will minimize the impact of this condition on on their life going forward…so it's emphasizing the seriousness, but at the same time trying to keep the worry levels down.”*  *“Maybe they would, depending on what happened, they may come into some kind of rehabilitation program. I think that's very important because it gives patients time to ask questions, to reflect, to consider the new status quo that they're in. Often it takes time to it it it it just takes it, you know you know that's probably the main thing giving patients your time you know that that's kind of difficult for myself. But you know we have a service which will provide that, and that's invaluable.”*  *“probably it is just sort of that that ability to spend time with these patients and through pure volume of work that we're that I’m doing at the moment it's it's difficult to to do that”*  *“Well there, there are no interactions. So so once the patient is referred until they're seen in clinic, and there's probably no interaction with the patients, once they're seen in clinic, then we will arrange some tests, perhaps, and then we would write the patient with the results of those tests, and the need for any further tests or review. What we're trying to do is have very much a one stop shop. So patients are seen, diagnosed, and treated or discharged.”* |
| CL Clinician. P Patient. Redacted text shown in []. | | | |

| **Theme 4: Systemic Challenges** | | | |
| --- | --- | --- | --- |
| **Subthemes** | **Description** | **References** | |
|  |  | **Patient Experience** | **Clinician Experience** |
| Access to Clinicians | Both clinicians and patients expressed an awareness of how the lack of access to clinicians is contributing to delayed and accurate diagnosis of heart disease. There were some differences in patient stories, as not everyone faced issues getting an appointment, although this was the majority’s experience. Meanwhile, clinicians provided different perspectives on how to deal with this, with some advocating for more staff, while others suggested patients needed to become more independent to deal with existing systemic issues and improve their outcomes. | *“The first thing I like to point out is that I've only seen the cardiologist once in 2 years, which is astounding, because I’m sure that everybody else here and other people I've spoken to would have received a far better level quality of service from their health service”*  *“I would say at the moment. It's lack of access to GPs.”*  *“Basically, needs to sort out why you’re not able to see a GP when you need to.”*  *“That's different surgeries, I suppose you know. I've moved to this one, at my previous surgery it was the same as this one. But going back, you know, 20 years, exactly what he's saying, but nowadays it's, you know, even when my children were young we had to move surgery to actually get someone to see them in the end.”*  *“But as I say, I have absolutely no problem with with our surgery and from a personal point of view, my daughter she is a GP. She goes in in the morning at 7, and quite often doesn't get away till about 9 o'clock at night, mainly because of paperwork but not because of people.”*  *“I mean it has to be good access to GPs”*  *“But certainly, I have no contact with my doctor,”*  *“They call them that they do most of the procedures on and the wait when for a follow up appointment was always jam packed with people. They just couldn't cope with the number of people they're saving.”* | *“From a doctor's point of view being seen quite promptly. So I you know especially with kind of everything at the moment isn't it like people feel they're waiting ages to be seen by a doctor, which you know, you can understand if you're feeling you're getting chest pain, and you're being told your next appointments three week’s time to even be seen. So I think probably like promptly being seen by a GP.”*  *“Lack of funding. Lack of sufficient doctors, nurses, and health workers in all aspects.”*  *“I think we need to empower a patient as much as possible and not leave them in a dependent state on the NHS because the NHS cannot, can no longer provide that sort of mollycoddling.”*  *“I think, up here in [City] it is a very, you know the nearest cardiologist is you know over several hundred miles away, so like we're quite isolated as well…So, you know we don't have a a cardiologist on the scene who can get like you know, cardiology range or consultant down to the department to to look at an ECG.”*  *“I can't be in a long-standing provision because I have more coming up the back that needs to be sorted,”*  *“I think that basically a lot of diagnosis is made in secondary care, so if a patient has an acute set of symptoms that goes to the ED department.”*  *“Don't start drugs going into a bank holiday, because I know the out of hours doesn't exist”*  *“Access to primary care itself, and the patient actually managing to make an appointment with me,”*  *“If there were more staff that we had, if there was if there, if it wasn’t the GP crisis if there was more GP surgeries if there was more people like myself working then we could get through more patients, there wouldn't be the delay ,there wouldn't be issues getting appointments, so I mean that's if that was solved, then I’m sure things would be better”* |
| Access to Investigations | Many clinicians described difficulties accessing investigations, including blood tests, ECGs, and CT scans, highlighting a key contributor to delayed diagnosis. Although patients did not explicitly express challenges accessing investigations, data from clinician interviews suggests this has a substantial impact on patient experience and patient outcomes, particularly in regard to efficiency of the heart disease diagnosis | *NA* | *“Actually being able to arrange some investigations,”*  *“The monitoring is a a very important part of what we have or should be doing. Regrettably, this is not something that is happening as frequently and as well as we would like it to happen”*  *“we work very hard in cardiology to make sure that GPs have open access to as many tests as they need, so that includes an ECG, heart tracing, a blood test called BMP and ECHO scan, ultrasound scan of the heart, and they can all be requested open access by the GP.”*  *“Once we’ve got hold of patients generally access to tests, it's not too bad, but some of our tests are sitting sort of four months, four months behind.”*  *“Availability of some tests. So I mean, although there's although there's there's waiting lists for certain things, you know, for example, we're in the in the highlands of Scotland, and we generally struggle to attract staff. We're quite lucky in our department but, for example, the Radiology Department, the X-ray department are understaffed and just last week we were told we've got no access to a certain test, CT scan for coronary disease for the next four months. So that's a real practical issue about getting access to tests.”*  *“It can be very difficult to if you've got a complex ECG, or complex patient to to get you know a good handle on what a potential diagnosis could be and also limited blood tests and investigations out of hours as well can be quite tricky to to yeah to get an accurate diagnosis.”*  *“although it’s different, you know, for you know for us the [Hospital-name] takes our cardiac ischemia ECG’s but for arrhythmias, they don't want to see those so that leaves us like in the lurch from that point of view. Obviously imaging as well, we don't have access to, you know media ECHO overnight or anything you know so uh we’re kind of stuck from that point of view”*  *“I think it's now more national, but we got access to proBNP quite early… we were well ahead of the curve and early adopters to the concept of 24 hour blood pressure monitoring”*  *“then with getting access to tests and investigations so again, even in their own practice, there can be a wait before you, or delay before you can get your bloods done, and we can obviously, in an urgent situation, get bloods and ECG done there and then, but normally it's a routine it might take a week or ten days.”*  *“The availability of investigations probably number one you know whether that be blood tests, stress tests, cardiology intervention, you know. I I suppose it's how that is done, and how quickly it can be done you know things are limited, you know, for the number of years, particularly where I work in Northern Ireland”* |
| Accessibility of Existing Systems | Patient experiences have revealed several concerns with the accessibility of existing systems, such as how to book an appointment with their GP or give the correct information when filling in an online patient form. These issues lead to a lack of patient trust and confidence in the healthcare system, which ultimately acts as a barrier to accurate and timely diagnosis. Clinician experience of these issues was also evident, although there was a general sense of powerlessness in the way clinicians expressed these concerns, suggesting these issues stem from greater systemic problems and contribute to both patient and clinician challenges in accurately and efficiently diagnosing heart disease. | *“But the group said, if you're in AF to just go to A&E. And the cardiologist wrote me a letter saying why, so they can actually capture the fact that you're in AF and what type of AF and you know the extent of the AF so it would make diagnosis and treatment easier, which I didn't know about. But also to ask for a copy, so you could take it to your doctor or if you had another event, you could take it with you. So I did all of those things”*  *“in NHS Scotland, which is obviously entirely independent of NHS in the rest of the United Kingdom and they classified a heart attack, or the myocardial infarction, it was a non-life threatening condition and you had to pay for your prescriptions. And yet, four years later I was diagnosed with diabetes, and doctor’s exact words were to me: I've got good news for you, you won't need to pay for your prescriptions anymore. So I I still fail to, how a heart attack is not a life threatening condition? Because I would imagine that, I’m no expert, but I think once you've had one, the potential to have another one, or problems related to your heart, would increase.”*  *“It’s access to your GP and you phone up on your first line you've got to get through to the receptionist which is really hard, and then they can you go through a you know kind of interrogation to find out how serious,”*  *“Unfortunately…in some cases the the surgery, and I know GPs personally, the surgery is adapted to suit the doctor not to suit the patient and it's very difficult these days…you get the opinion that it's organized for to suit the doctor. The patient is just an encumbrance to the daily daily lives of everybody connected with the surgery. And unfortunately this is the problem with the NHS in total, it's run by incompetent managers. Unfortunately, the staff are doing their job, and they're doing it to the best of their ability. But unfortunately, there don't seem to be anybody looking after it in total. It just seems to be utter chaos, and it's it's a it's a great pity because the National Health Service is a fantastic organization, if it was run right, but it's not. unfortunately. But that's just my opinion.”*  *“Yeah, I think having a phone line even for someone like me… to refer yourself through, like counselling, or in just basic primary mental health. And there's no, there's a phone number, but I mean I can try calling it, but will anybody answer? And I was just trying to explain to somebody else that I can't yeah, I can't explain what I'm going through, even for me and I’m but if I’m feeling rubbish. And it's some, that adds to all of these barriers as people have talked about, they definitely add to the stress and health issue and you know with things escalating”*  *“I said, I want to go home. You can't. What do I have to do to go home? They said, well, you've got to sit a thing called a Mocker Test. It was a cognitive test and get 84%. I said come on then, she said no, you’re not ready. Well, anyway, I got 86%. And I said in that case, I presume I can go home, and I knew it was far too early. The hospital staff weren't very happy”*  *“I act as a carer for my wife who’s had MS for 40 years. She’s paralysed. We, needs my help with every daily the activity from getting out of bed to getting back into bed yet there was absolutely no, and this was known, this was all recorded at the hospital, there was no, there was no help or assistance offered in that direction whatsoever.”*  *“When I was in a hospital, that was brilliant. I was given my test results but afterwards it was very, very difficult to get any communication with [Hospital- name] because they were just too busy. In [Hospital- name] they got 4 labs, they call them that they do most of the procedures on and the wait when for a follow up appointment was always jam packed with people.”* | *“I guess that isn't the model in general practice, unfortunately, that they may well see doctors in the beginning, and then they're probably seeing health care assistant who might take their bloods and blood pressure, and then they might see a nurse who goes through those results with them, and I mean, even if they they are being followed up in clinic, they're often not followed up by the same consultant that they saw initially.”*  *“The NHS does still kind of use the the mail basically, so and actually, we don't routinely kind of have an email back within a couple of days from a clinic as to what's happening”*  *“I think it's just a big organization isn't it like the whole of the NHS, and that's probably quite uniform across big organizations, but the poor patient stuck in the middle of it, is kind of trying to speak to you, and we are saying it is not us, it’s the hospital. They then speak to someone secretary at the hospital who says it's in progress, or, as far as we know it's in the post.”*  *“what is life-saving usually gets done, but life changing procedures, experiences and treatments simply because there is no capacity to see all these people so quickly.”*  *“The problem there is, because vascular services are centralised, not every hospital will have vascular services, so they may be in another hospital, and they will get transferred to the Hub or the main hospital with the with the vascular centre. So for the ones that come to straight to A&E, so the more acute or urgent presentations, they more often than not, unless it's a complete I guess misjudgement, will be admitted to be managed as an inpatient.”*  *“patients coming who clearly don't speak the language, or for whatever reason cannot communicate and we've not been made aware of that. So either the referrer hasn't told us or we've not, either I don't know, found a way of getting help, or indeed, even trying to book a longer slot”*  *“a problem getting data out of our archaic NHS systems is quite challenging in terms of how well, how well are we actually doing so”*  *“Well there, there are no interactions. So so once the patient is referred until they're seen in clinic, and there's probably no interaction with the patients, once they're seen in clinic, then we will arrange some tests, perhaps, and then we would write the patient with the results of those tests, and the need for any further tests or review. What we're trying to do is have very much a one stop shop. So patients are seen, diagnosed, and treated or discharged.”*  *“I think it's difficult to when we are in a cash limited and manpower limited service, I mean, there's a it's a restricted service to to what patients might like, and we try very hard to be efficient in terms of seeing patients just once in the clinic and then discharging them. So it's very much a one stop shop. Make a diagnosis. Here's a management plan.”*  *“all of the practices now or all the pharmacists, the community pharmacists have reached their threshold for nomad systems or deliveries”*  *“they have very, very, very, very very very poor information given in secondary care.”*  *“I don't start drugs going into a bank holiday, because I know the out of hours doesn't exist”*  *“We have we have a limited resource, and unfortunately we have practices closing and and we're taking on few patients from surrounding practices. So you know our pressures are are great, patients can’t get slots and that that's a that's a big issue…people were phoning multiple times but you know the demand is is enormous on the service”* |
| Approach to Medication | Several patients described feelings of confusion and uncertainty with the way their medication was being managed, with specific concerns that drug-based treatment was an ‘easy way out’ for the health service to discharge them as quickly as possible. The lack of follow up on medication type or dosage left many patients feeling unsure of what they were taking and why, and how this was affecting their health. This approach to medication could contribute to a distrust of medical professionals and elicit fear in patients that reaching out to their doctor will result in further prescriptions, encouraging a pattern of avoidance behaviour. | *“had about five or six years with just medication”*  *“But they I've I've always complained about the amount of medication I’m on because I've never had an incident. But they they, they, they say, to me in in their defence if you want to phraseology it that way, that is it because of the medication I'm taking that I've got good readings or have I improved? But they don't want to stop a medication in case it is the medication that's keeping me on a straight and level path, so you can't win, really, you're stuck between the rock and a hard place”*  *“I’m just recovering from a bout of pneumonia and I went, I lost quite a bit of weight, and I went to the doctors, and he said you know you're on far too many medications. We're trying to kill you with kindness. So they've actually taken, stopped two of them. So why couldn't they have stopped the 2 of them before?”*  *“But I haven't had any treatment. I'm. I'm just taking drugs. They they they they they discharged me with a bag of drugs, and I'm still taking the drugs 2 years later.”*  *“I had a a nurse that would ring me. She rang three times. And it was only after a while I realized that her sole objective was to try to get me to take more drugs. That was all she wanted to do. I said, I'm taking loads of drugs. How many more do you want me to take?”*  *“I too, have also been told to do this by my doctors. Not once do they ever ask me to go into the surgery to do, check my blood pressure. I have to ring once a year with my blood pressure results over, done over three days at the same time. I'm on repeat prescriptions for all my medication for my heart conditions and and no one questions. No one questions.”*  *“At first, they they put me on loads of medications, some of them making me more ill. So, I had to stop them by myself. Then I was told to keep only one, bisoprolol. Even then I was getting chest pain from that side effects. I don't know what it is”*  *“I had a single stent fitted and other than being on all the tablets that you can imagine God created to do with heart conditions.”*  *“I'm stuck on Warfarin as one of the pills so that means I’ve got to be tested about once every 3, 4 weeks, which is not so much fun,”*  *“I know that when I went in to the doctors for the first time to be checked over and they discovered the erratic heart beats and all the rest of it, they gave me a medication of of warfarin which was based on my weight. Now my weight then was quite high. It was about 24 stone. It's now down to about 13 and a half stone, which is quite good after about 10 years or so, but I'm still on the same level of warfarin which sort of beggars disbelief somewhere along the line because in theory well, I’m not quite sure how it's supposed to work and I, when I asked the surgery, they weren't quite sure either. So, it seems to be Sod’s law. If it works, keep going and if anything changes, the whole thing will change. It seems a bit ridiculous to me.”*  *“I went yesterday to have my bloods taken for my annual diabetic review so if by chance one of the many tablets I take is uh, I have the wrong dosage, how are they ever going to know unless I’m specifically going in to see them about symptoms that might be caused by that?”* | NA |
| Communication within the NHS | Poor communication between different services in the NHS has been brought up as a key concern by patients, with many describing experiences of their information being lost or misinterpreted during transmission. This can contribute to delays in diagnosis, as well as unnecessary emotional stress for the patient and resulting worse outcomes. Meanwhile, clinicians describe a range of experiences with communication between services, with some praising the existing systems for efficiency and ease of use, while others describe difficulties receiving adequate and accurate information from other departments and clinicians. | *“But, there are occasions where I'd go along, and they would have carried out a test and then lost the results, so much of it was pretty pointless”*  *“I think that's you know the continuity of care is reliant upon very good recording by GPs, which I think doesn't always happen.”*  *“Bizarrely our local Trust, one of the departments there, seem to be so unconvinced that the GP use the information correctly, is that they've taken to writing the follow up letter that would normally go directly to the GP, they write the letter to the patient with a copy to the GP. Presumably, so that the patient knows what questions to ask when they go to visit the GP rather than relying on the information being available on the screen, and then proactively dealing with it. Oh, that's that's a dreadful scenario isn't it? When when somebody some consultant is so concerned that the GP won't follow up that they make sure that the patient has the information. But already every department in our local Trust sends a copy of every GP letter to the patient. But this one department, whether it's a trial or whatever, is doing it the other way around, they’re sending the letter instead of to the GP with the copy to the patient, they're sending the letter to the patient with a copy to the GP.”*  *“I was a bit horrified at the whole thing. I still believe that most of the hospitals have not caught up with the technology to transfer information from one part of the hospital to another part of the hospital and I think that’s a little bit ridiculous all the way round.”*  *“I’ve touched on it before the the communication between the the nurses and the doctors and the surgeon was sometimes lacking. For example, I was prescribed, post operation, I was prescribed a certain drug for which is normally used for lowering blood pressure but it was more to do with protecting my heart after the surgery and the nurse couldn't understand why I was I was prescribed that so they they didn't give me the dosage for several nights. It was only when I raised it with the cardiologist who came to visit me on his round, he was not happy at all, and that's that relates to communications. The complete lack of communication between the medical staff, the nurses in this case and the senior doctors and cardiologists. And surgeon, sorry. So that's that's a major issue, I think, in the NHS.”* | *“I I mean it is like often I think they're quite frustrated by it in all honesty. So you know we'll do a referral whatever, but it can take a while, you know, if they're seen either in the hospital by the time that letter is typed up and sent back, because the NHS does still kind of use the the mail basically, so and actually, we don't routinely kind of have an email back within a couple of days from a clinic as to what's happening, so I think they're quite frustrated, because often they'll book in to see a doctor and they'll say, oh, I saw someone two weeks ago, and that letter, because of all the backlogs hasn't been typed up and arrived or you've had one bit of paper with the medication they want prescribing, and then they said, oh, you know I'm supposed to come in in a couple of weeks to see whether it needs changing, but we haven't yet had that information through. So I think to be honest, a lot of them are quite frustrated, you know, and you can understand why, because from their point of view, they think, why, why, in this day and age isn't it an email just sent, and that's available? I suppose they, you know, they kind of they don't understand necessarily the process of what's happening. Where we are there is quite, they generally get hospital appointments, but there is frustration that often they are cancelled, and they are sent letters in the post, or they’re rung up and told their appointments are cancelled before they've actually even got one through the post. So loads of them will say I didn't know I had an appointment, but someone rang me and told me it was cancelled.”*  *“Patients coming who clearly don't speak the language, or for whatever reason cannot communicate and we've not been made aware of that. So either the referrer hasn't told us”*  *“GP will contact me using a system called clinical dialogue. It's a bit like email, but it actually automatically uploads to the electronic patient record and that makes it quite helpful. It's less helpful because it's just it's it's one to one, so we can't actually involve other health care professionals in that discussion. So it's not quite so good as email, but it is from a governance point of view, excellent, and often the GP will send us a request for some advice we'll give the advice, and that's the end of it.”*  *“So we would expect the GPs to put that in the letter, and to give us the information. So that's going to be things like hopefully, heart rate, blood pressure, ECG, any symptoms of chest pain, or breathlessness, or swollen ankles, and including their kind of findings when they listen with a stethoscope and we would expect that to be put in a written, the written referral on Clinical Dialogue, or through the Sky Gateway referral.”*  *“So we have this thing called RTT, I don't know if its Scottish or whether it's UK, and it's supposed to look at when the patients were referred, and when they were treated. The problem is that are none of our clinical systems can actually join up all the the bits of the pathway. So actually, nobody has a clue. None of us. I do not, I have no idea how long it takes between referral to treatment of our patients. So that that's quite frustrating when it comes to service redesign, or service delivery”*  *‘I think if we had better reporting, I think if we had better standard setting and reporting of outcomes that would give us more clout if you like to to make a difference.”*  *“I think paramount is just being a working as a good team, a good clinician working well with you know the other doctors, the nurses, making good like, the radiographers, making sure that you know that they can see that you know every effort’s being made to to try and get the diagnosis, and follow treatment and from there, with the you know clinicians you know, and in sort of tertiary centres for us, and you know they have the retrieval team and helicopter team, and all these things.”*  *“One of the problems with that is, is that complex cardiac conditions are managed in ED; that’s fair enough from a safety point of view, but ED has no mechanism to give us feedback, and there's often these things, these wee scraps of paper saying, you know, has been referred to cardiology, and it hasn't happened.”*  *“We've gotten to be little lazy about that stuff, but locally recently there's been fewer practice-based pharmacists so that’s allowed me to realize I can't really rely on them. So I've kind of come back to developing strategies for myself.”*  *“When they come back I suppose the interface between ourselves and and and secondary care, sometimes not always as it as it could be you know there things are happening that maybe we are not aware of, or how we could communicate with them could be better.”*  *“Well, if say, if someone's been seen like a patient might have been seen, you know, on the Monday by you the sort of the the team we have is called Our Hearts Our Minds, and it's a nurse led service, together with a cardiology back up, but the patient may be seen on Monday, and then they come or contact the practice on Tuesday or Wednesday to to discuss what's happened and really I I don't know anything about what's happened, and there's no communication to that point, and I obviously it’s only been a short time. But sometimes it's it's even longer, and you're kind of in the dark, and it's It's it's not very good you know, maybe maybe they'll get way of just getting that information immediately.”* |
| COVID-19 Aftermath | Both patient and clinician experiences suggest that the recent COVID-19 pandemic has amplified existing systemic issues related to access to clinicians and contributed to delays in receiving an accurate diagnosis. While patient experiences post-pandemic have been mostly negative, some clinician views support the shift towards a more efficient way of dealing with certain patient interactions, such as implementing telephone appointments instead of doing everything face to face. | *“I was in dialogue with my GP practice. But that sort of ground to a halt during the pandemic for some reason, that they you know they they weren't following up, asking for my readings, so I just stopped doing it”*  *“They put me on statins for cholesterol, and blood pressure tablets, aspirin all sorts of things, and for the first year I had to go every 3 months for blood tests and that then went to 6 months and since the pandemic it can just be whenever I suppose they've got a space.”*  *“I would say to my GP, pre-Covid because now you you can't get to see them obviously as easily,”*  *“Whether it's the pandemic or not, I've had since two years I've been back, no one has been in contact with me from the NHS.”* | *“What happened during the pandemic of COVID-19 has taught us a fantastic example how suddenly we are able and have become comfortable deciding to contain a follow up clinic in a telephone conversation.”*  *“During COVID we found this because we thought, can we make use of some of these things? But what a lot of the patients said was missing actually was just not, the the sort of more direct contact. ”*  *“I mean right now in the kind of post-Covid era, patients have a lot of problems getting access to a GP so I mean, that's a big barrier right now, and we have we have a limited resource, and unfortunately we have practices closing and and we're taking on few patients from surrounding practices.”*  *“Patients can’t get slots and that that's a that's a big issue, you know, I think, there’s probably a huge, undiagnosed burden from Covid so we're trying to catch up with that and those patients may be not people who will determinedly approach us for review they may be people who would maybe sit back, and maybe their symptoms, they potentially feel they’re not that serious, but in reality they're probably the potentially the sickest people so so it's it's difficulty in accessing yourselves,”* |
| Geographic Factors | There appeared to be a real case of ‘postcode lottery’ when it came to the quality of treatment patients received and clinicians were able to provide. Both patients and clinicians referred to how their geographic location contributed to patient outcomes, specifically when it came to the efficiency of diagnosis as a result of access to resources. Clinicians who worked in more remote locations in the UK were more likely to express issues with having access to staff and investigations, suggesting difficulties reaching appropriate diagnoses and treatment outcomes for their patients. | *“The local hospital was about a mile and a half from where we were, so they decided to basically throw me in the back of a car and bring me to the hospital.”*  *“I can only think it's a bad, [city] is a bad area.”*  *“In [local-city] they said, we have the option to go to one of the [distant-city] hospitals because they've got spaces or stay in [local-city]”*  *“I'm very, very lucky, because I live well 15 minutes if you're being blue lighted, but normally about half an hour from [Hospital-name] which is obviously the best heart hospital in probably in Europe nearly so, I was very lucky,”*  *“I'm lucky because I because of where I live.”*  *“There’s a sort of a minor injury unit in my area, and so I I see people plodding off down there to get things like antibiotics.”*  *“I’ve had no real problems with the Health Service where I am.”*  *“I don't think it's the same case in Wales”* | *“We'd normally take a history and examine them to see what we could find on clinical examination and I guess, depending on that in the GP surgery we could do an ECG, like a heart tracing for them, blood tests, and then probably send them off to have more investigations done through the hospital. So we tend to organize them in terms of like a 24 hour ECG, so 24 hour tracing of the heart we can do. We can do blood tests, have a look for heart failure. Where we are, we can order Echos, so like the scans of the heart, to have a look at them.”*  *“A barrier could be, could be, the lack of experience, could be the lack of facilities in the hospital or the clinic,”*  *“We're in the in the highlands of Scotland, and we generally struggle to attract staff. We're quite lucky in our department but, for example, the Radiology Department, the X-ray department are understaffed and just last week we were told we've got no access to a certain test, CT scan for coronary disease for the next four months. So that's a real practical issue about getting access to tests.”*  *“So in terms of making the diagnosis, you know, you can make a you know, diagnosis of a an MI, and you know and you know theoretically they should go for angiogram but when they're 97 and  you know they have lots of co-morbidities and you know they they don't really want to leave the island than these things can be, play an important role as well. ”*  *“I think, up here in [City] it is a very, you know the nearest cardiologist is you know over several hundred miles away, so like we're quite isolated as well. So it can be very difficult to if you've got a complex ECG, or complex patient to to get you know a good handle on what a potential diagnosis could be and also limited blood tests and investigations out of hours as well can be quite tricky to to yeah to get an accurate diagnosis. ”*  *“We are very isolated. So, you know we don't have a a cardiologist on the scene who can get like you know, cardiology range or consultant down to the department to to look at an ECG”* |
| Limitations of Existing Diagnosis Protocol | There appeared to be a general frustration with how current procedures for diagnosis operated, both from clinicians and patients. The existing system seems to be lacking in flexibility, holistic considerations, and general accessibility. Some patients expressed concern for how they were not taken seriously because they did not fit the exact mould of certain heart disease diagnoses and how this contributed to delays in their diagnosis or resulted in poorer health outcomes. Moreover, clinicians echoed these concerns, describing their own personal frustrations with not being able to go outside of the strict protocols to diagnose unusual symptom presentations. | *“I just remember at the time being surprised how inexact the way the diagnosed was, that they were relying on a 24 hour period of time to decide what it what it was, or whatever. If I didn't have an episode, there would be nothing, nothing to gather, and, as it happened, there was barely a blip in the 24- or 48-hour period, but they still diagnosed paroxysmal atrial fibrillation.”*  *“they would observe me for a few hours and discharge me. I think I went to hospital twice, and on the third occasion, I thought, well, what's the point they are just going to discharge me so I didn't bother.”*  *“I have no way of knowing my cholesterol unless I go in for a blood test.”*  *“I purchased myself like one of these blood pressure monitors for at home, and I used to take it almost daily, and the readings were within normal bounds. But whenever I went to the doctor’s it was never under 160 over 92 or something and I discovered white coat syndrome at that juncture and I pointed it out to him. So one of the nurses, the the nurse practitioner she tried me on both the the blood pressure monitor, the electric one and on the hand held one. And whenever she did it on the hand held one, it was about 25 points lower and she did it within within 10 minutes.”*  *“I asked if I would be monitored in some way, and I was told only if you get symptoms. I didn't have any symptoms.”*  *“There needs to be, I think, better monitoring of people with no risk factors and also better information given to them, like when I was diagnosed with diabetes, as I said, it was two years, two, three years before, three years before I got my stenting done, there was no discussion, I I mean, I now now, I kind of knew, anyway, but I know that most people probably wouldn't, I know now, there's a huge connection between heart disease and diabetes, but it was never discussed you know, with all the different people I saw when when I and I did, the DESMOND… I don't remember anything about heart disease being discussed and yet the two are very closely connected. So I think there needs to be better information given to people who have conditions that are very likely to lead to heart disease.”*  *“I think I've already said that that's been the major problem. In fact, I think*  *my problem also associated with pre-care as far as the NHS were concerned”*  *“I gave up on NHS to be honest. I thought it was only me.”*  *“One of the consultants told me that I don't look like a heart problem because I am still young, and I don't, I'm not overweight, or I don't drink or alcohol anything like that. So I shouldn't have a heart problem basically. So I was like, oh, okay, so they discharged me two years ago.”*  *“But I've never had a problem with cholesterol, whereas I think the NHS is totally focused on what your cholesterol level is.”*  *“As I learnt in Cyprus, cholesterol level to a Diabetic type one is not what causes the arteries get clogged up, it's it's a a purely diabetic problem, and it's and there is no focus on this in the NHS...That's something that that from how he explained that happened there is very much lacking in in the NHS, because they focused only on my Type One diabetes, tried to look after that but weren't really concerned with all of the, what I now know, are very considerable and frequent side effects and really that's why they have to spend a lot of money when it all goes wrong,”*  *“when you visit the do visit the doctor, you are forever repeating yourself you're having to give your diagnosis again. What's happened to you over the years because they don't know anything about you. And basically, even though the information is there in your file on the screen they are still asking you your history and I find that extremely frustrating. I don't know if others feel the same about this. I know doctors see many people, but when you make appointments surely they read your records before you go in to see them. You know your bit of history. Well, I I find that this doesn't happen every time.”*  *“It seems to be standard procedure that they don't give you any any any of this data or information like test results. If that was something I would really like to have in my position, so I can get some idea as well. But you. You have to request it. It's not automatically given to you. I think that should be something that should change.”*  *“They just couldn't cope with the number of people they're saving.”*  *“You know the the classic case of the the operation was an unmitigated success, but unfortunately the patient died.”* | *“In terms of, it's quite variable, really. So say they had an irregular heartbeat, lots of people have, ECG monitoring, but actually we probably don't do loads in the community to monitor them while they're being seen. yeah, that's probably, yeah, probably about it for monitoring, really.”*  *“So I guess an ECG heart tracing which we can do, although that's a bit variable, because it's only a snapshot of kind of 10 seconds you're looking at really,”*  *“what is life-saving usually gets done, but life changing procedures, experiences and treatments simply because there is no capacity to see all these people so quickly. ”*  *“we find that's quite inaccurate in terms of people's estimate, so we do. So we may do at the start a walking test, the treadmill test, and then we'll have an objective assessment of how long and what's the distance they walk before the symptoms come on. But of course that is in the setting of the laboratory so when you walk outside a lot of things vary right? Whether you're walking up the slope, down a slope. Is it windy? Is it cold that day? Are they carrying stuff? And, at the end of the day those are the things that are relevant because it's related to what they need to do in their daily lives. So the objective measurement in the what we call the vascular studies lab is useful, especially if you're planning intervention. So you have a and then you can or you, you know you might be useful to compare the later time, whether it's improved or not.”*  *“The sort of observations really. I guess that's a sort of, I guess, if you mean numeric. The troponin, because we’re so in the acute stage, you know sometimes you know, a negative troponin doesn't necessarily mean that you are still not sort of, still don't think this is an MI, you know, in a lot of cases, you know I’ve seen patients with negative troponins, and then then it's been raised later and they’ve had you know definitive MI so yeah, like the blood test sometimes is not you know they help and judge, but I wouldn't put any definitive diagnosis sometimes directly on the on just one blood test”*  *“I was really kind of, came to the practice about ten years ago, and I said, folks, I think we need to be doing 24 hour blood pressure monitors, not treating people based on one-off readings”*  *“I've had about I can easily say about five patients that were admitted to a surg-, a medical ward and started on blood pressure medication based on blood pressures that don’t even make NICE guidelines and I bring them, they come out to me, and I’m like I don't - can we just put them on a bloody monitor, and then itt's normal. And when you spend five minutes interrogating them, they’ve just got a a death or a grief or something that’s happened.”*  *“Then you have the getting into the hospital investigation or secondary care investigation obstacles too, they they follow quite strict guidelines, which I find a little bit of an obstacle because as a clinician, I may well feel very strongly that this is a ischaemic heart disease and and you make that case in your in your referral, but if it doesn't meet the criteria, and then it is rejected.”* |
| Time  (Efficiency, Time Restrictions, Waiting Times and Delays) | Time was one of the most discussed aspects of diagnosis, including both praise for the efficiency of diagnosis and treatment, and frustrations with time restrictions in appointments and long waiting times for appointments and surgeries. This was a substantial subtheme amongst both patient and clinician data highlighting how this is a great systemic issue that contributes to existing challenges in providing patients with an accurate diagnosis and positive treatment outcomes. The benefits of efficiency can also be seen in our data, as patients who described a smooth and quick diagnosis process reported better health outcomes and higher satisfaction. | *Efficiency*  *“Very quickly I was diagnosed with a particular arrhythmia called arrhythmogenic right ventricular cardiomyopathy”*  *“The ambulance came, and they wired me up and everything and their initial diagnosis was paroxysmal atrial fibrillation or occasional atrial fibrillation, and I was just having a significant episode, then to the local A&E, who again said, yeah, it looks like you you've got AF but they said we'll let you home, if it, if it goes back to sinus rhythm, and which it did and then packed me off to the GP. I went to the GP. I’d had one or two little occasions in between then, from when I'd been hospital in hospital to the GP, when I thought, hmm was that the same thing again but they only lasted very short period of time. With the GP I then she said, I I need to refer you to a cardiologist,”*  *“When I was discharged from the hospital, they gave me a bag with some medications in it and a letter from my family GP. The following day, a family member took the letter down to the surgery and the following day after that, I got a phone call asking me to attend at the surgery later that afternoon.”*  *“It was a very, very precise definite diagnosis and a very precise, definite solution all based off that one figure of a super-fast, so a tachycardia, and the solution was equally as clear-cut and simple.”*  *“I just took myself to to the GP and they sent me straight to specialists the next day or so.”*  *“the ambulance came in 20 minutes and an ECG told me straight away. Yes, you're having a heart attack”*  *“I never had a problem, we do a thing called MyGP in the morning. You get a phone call from usually a receptionist and by mid-afternoon you've probably seen the GP if you need to. I've absolutely no problem with that.”*  *“I had a GP who recognized very quickly that the symptoms I was getting were most probably to do with heart disease. I was referred very quickly to a cardiologist. The cardiologist referred me very quickly to have an angiogram to have it looked at so for me all happened the way it should happen.”*  *“When I spoke about a referral to mental health. I was seen quite quickly, and had and have a series of CBT and stuff very quickly where we worked at, and my anxiety about going away, I mean, like I said, I've been away for six weeks, so my anxiety about going away went away actually quickly and I think that was because I had intervention quickly. I had somebody to help me quick, so it's the speed of, the speed of response, I think is the thing.”*  *“I was unconscious for three days. They had to put an angio-, an aortic pump in, took it out after three days, and decided that I needed a new valve, within a week I was in surgery, having a new valve, and they did a bypass as well. Everything it just happened so perfectly well and I have no complaints”*  *“I can't fault them when I was having the event, as I was, rushed straight to hospital and had 2 stents fitted within a couple of hours”*  *“I know in my case, after I got identified I was given a chance to have my heartbeat restarted again, and that was quite an interesting experience. But again very quick, very efficient.”*  *“Doctor says I will write to your GP today the letter will be dictated today. They always say this. Yes, they dictate the letter. It then goes by pigeon post to the typist, 20 feet away, who sit and look at it for 3 weeks. They then type it, print it out, send it back to the doctor, and something like 6 to 7 weeks later the thing gets sent out.”*  *Time Restrictions*  *“I probably didn't get the time that I felt like that I needed”*  *Waiting Times and Delays*  *“the cardiologist just said, well, we can fit a device that can help you, two years later I'm still waiting for it...* *for two years now, they have been saying that they want to implant a small device in my chest but they are talking about it and not doing it.”*  *“I am still waiting for an operation.”*  *“We went through a process where we were waiting quite a long time for an ablation, even though it'd been we'd been put on a waiting list,”*  *“we have the option to go to one of the [distant-city] hospitals because they've got spaces or stay in [local-city] and although I would have liked to have had it done quicker, I kind of thought for my family that would be dreadful if anything happened or you know, if my husband had to come, he'd be on his own, he’d have no support. I knew I’d be in possibly overnight, so I elected to wait, and as it turned out, it wasn't as long a wait as they anticipated, but I didn't look on it unfavourably.”*  “*She told me there was a two hour wait for the ambulance.”*  *“I asked for the recording. I had to wait three months, which didn't help”*  *“then you have to book in three weeks in the future that you're going to be ill, and by then it's too late, and the the alternative is to go to A&E and sit there for seven hours which you know obviously we're going on to more political things by going further.”*  *“this is actually getting worse because I I remember some of my family they, you know, it might take 10 days to get a a a telephone appointment with the doctor, even if it's a a pretty acute condition”*  *“That's all they said, I'll ring an ambulance and the ambulance will come out to you. Four hours I was in the ambulance and 26 hours I was in the corridor on the trolley. And since then nobody's been interested in me.”* | *Efficiency*  *“we don't have great issue in getting in getting simple baseline investigation done, we can really do them, you know, immediately so I don't have a problem in that way.”*  *Time Restrictions*  *“Facilities will help, you know, tools to help with the communications like drawings, pictures, things that would be able to help, which can inform the patient better about the condition and then having enough time.”*  *“I think time is a challenge in the NHS. You know, in this case I'm only given 10/15 minutes to see one patient, which I don't think is enough, you know, giving, you know we need to have a you know, some conditions, especially in the complex ones will require you know a long, you know, half an hour, an hour time to communicate properly with the patient.”*  *“We don't routinely do quality of life questionnaires in the clinic, partly because of time, but also there are problems using some of these tools as well.”*  *“understanding, because you have a very short time to understand the patient, their concerns, their daily lives, and their expectations and hopes and dream isn't it, is all within that so we have quite a short time to do that.”*  *“so communication issues, and you know we do have interp so say language. We may have interpreters but very often that is, you know, there are still limitations with that. And so, yeah. So communication in an already limited or timed setting.”*  *“patients coming who clearly don't speak the language, or for whatever reason cannot communicate and we've not been made aware of that. So either the referrer hasn't told us or we've not, either I don't know, found a way of getting help, or indeed, even trying to book a longer slot”*  *“I think some things are quite simple and used to be done and could just be having more time and building up a relationship. And I think a lot of that is lost in modern medicine...So, for example, again, belief that, you know there might be a reason why they are not opening up and telling us everything. Some of these would have been explored already. So by the time they come here they understand that we're trying to help. Not so, there's no point, you know, we're not judging, for example or and now not only do they not have necessarily a a a doctor that knows them well, the doctors, the GPs, don't have time to do this”*  *“it's a restricted service to to what patients might like, and we try very hard to be efficient in terms of seeing patients just once in the clinic and then discharging them. So it's very much a one stop shop. Make a diagnosis. Here's a management plan”*  *“it's a restricted service to to what patients might like, and we try very hard to be efficient in terms of seeing patients just once in the clinic and then discharging them. So it's very much a one stop shop. Make a diagnosis. Here's a management plan”*  *“He was diagnosed with unstable angina, sent to the Cath Lab, got Cath-ed and got four stents and was discharged within 24 hours. He had extreme trauma after that. He made an appointment to come into me, and he said, oh, he called me Dr [CL010], that's what they do, I have 16 questions, and I had to lean into that in a 10 minute consultation.”*  *“Second obstacle is is time, access to primary care itself, and the patient actually managing to make an appointment with me”*  *“Maybe they would, depending on what happened, they may come into some kind of rehabilitation program. I think that's very important because it gives patients time to ask questions, to reflect, to consider the new status quo that they're in. Often it takes time to it it it it just takes it, you know you know that's probably the main thing giving patients your time you know that that's kind of difficult for myself. But you know we have a service which will provide that, and that's invaluable.”*  *“probably it is just sort of that that ability to spend time with these patients and through pure volume of work that we're that I’m doing at the moment it's it's difficult to to do that”*  *“I would like to spend longer with these patients, especially in the initial assessment but we're maybe not making I mean I might, I might have suggest what the diagnosis might be before, but at this point they probably haven't got investigations done as so but you know that that's probably a limitation how how long I can spend”*  *Waiting Times and Delays*  *“From a doctor's point of view being seen quite promptly. So I you know especially with kind of everything at the moment isn't it like people feel they're waiting ages to be seen by a doctor, which you know, you can understand if you're feeling you're getting chest pain, and you're being told your next appointments three week’s time to even be seen. So I think probably like promptly being seen by a GP.”*  *“Actually, you refer to the local hospital, but they can here still be waiting kind of the waiting times are going up and up and up, and it could easily be six months before they are actually seen by any kind of specialist. So I think if we could have that prompter for patients, they would actually kind of feel more reassured by that”*  *“* *we'll do a referral whatever, but it can take a while”*  *“What is life-saving usually gets done, but life changing procedures, experiences and treatments simply because there is no capacity to see all these people so quickly. ”*  *“* *I think timely diagnosis is really key. So I think that patients waiting for three months, well patients waiting any time, it's just nonsense, and doesn't do anybody any good. So I think there's some people whose symptoms will get better, so time time can sometimes add clarity to the diagnosis, but generally, delays are just bad for everybody”*  *“I think time to diagnosis is important”*  *”Once we’ve got hold of patients generally access to tests, it's not too bad, but some of our tests are sitting sort of four months, four months behind.”*  *“there's waiting lists for certain things, you know, for example, we're in the in the highlands of Scotland, and we generally struggle to attract staff.”*  *“our literally, you know, absolutely obscene waiting times”*  *“the majority, an awful lot of our patients are managed in the emergency department, and there's three day waiting lists currently for the emergency department.”*  *“then with getting access to tests and investigations so again, even in their own practice, there can be a wait before you, or delay before you can get your bloods done, and we can obviously, in an urgent situation, get bloods and ECG done there and then, but normally it's a routine it might take a week or ten days.”*  *“that's probably a limitation how how long I can spend, and then I suppose, the the waiting lists, and and how quickly we can get people seen by secondary care and the cardiology team,”*  *“if it wasn’t the GP crisis if there was more GP surgeries if there was more people like myself working then we could get through more patients, there wouldn't be the delay”* |
| CL Clinician. P Patient. Redacted text shown in []. | | | |
